# Household characteristics associated with environmentally persistent free radicals in house dust in two Australian locations

**DOI:** 10.1101/2023.10.22.23297367

**Authors:** Wen R Lee, Prakash Dangal, Stephania Cormier, Slawo Lomnicki, Peter D Sly, Dwan Vilcins

**Author notes:** **Corresponding author:** Wen Ray Lee | |.

## Abstract

The association between air pollution and adverse health outcomes has been extensively studied, and while oxidative stress in likely to be involved, the underlying mechanism(s) remain unclear. Recent studies propose environmentally persistent free radicals (EPFRs) as the missing connection between air pollution and detrimental health impacts. However, the indoor environment is rarely considered in EPFR research. We measured EPFRs in household dust from two locations in Australia and investigated household characteristics associated with EPFRs. Random forest models were built to identify important household characteristics through variable importance plots and the associations were analysed using Spearman’s rho test. We found that age of house, type of garage, house outer wall material, heating method used in home, frequency of extractor fan use when cooking, traffic related air pollution, frequency of cleaning and major house renovation were important household characteristics associated with EPFRs in Australian homes. The direction of association between household characteristics and EPFRs differ between the locations. Hence, further research is warranted to determine the generalisability of our results.

## Introduction

The association between air pollution and adverse health outcomes has been long established, but the causal mechanism(s) behind this association are still uncertain (Kelly 2003, Kelly & Fussell 2015, Mudway et al. 2020, Risom et al. 2005). Recent studies propose a role for environmentally persistent free radicals (EPFRs) as a potential inducer of oxidative stress (OS), which may be the missing connection between air pollution and detrimental health impacts (Dugas et al. 2016, Gehling & Dellinger 2013, Guan et al. 2021, Pan et al. 2019, Ruan et al. 2019, Sly et al. 2019, Vejerano et al. 2018, Xu et al. 2021, Xu et al. 2020, Zhao et al. 2021).

EPFRs are formed during combustion processes and are present on small particulate matter (PM) with a mass median aerodynamic diameter of 2.5 micrometres or less (PM_2.5_), usually generated from traffic-related air pollution (TRAP), residential activities, industrial burning, and cigarette smoking (Chen et al. 2019, Gehling & Dellinger 2013, Sly et al. 2019, Vejerano et al. 2018, Wang et al. 2020, Xu et al. 2021, Zhao et al. 2021). EPFRs are particularly concerning as they are free radicals that can persist in the environment and in biological systems for prolonged periods of time (Pan et al. 2019, Sly et al. 2019, Vejerano et al. 2018, Xu et al. 2021, Zhao et al. 2021). Typically, a free radical has a short lifetime of a few picoseconds due to its highly unstable nature of the unpaired electron/s in the outermost orbit (Pan et al. 2019, Vejerano et al. 2018); however, EPFRs are a long-lasting stable compound as its valence electrons are continually cycling through pairing and unpairing by the transfer of electrons from organic particles to the surface of redox-active transition metals on particulate matter (PM) (Gehling & Dellinger 2013, Guan et al. 2021). These stabilised EFPRs have long lifetimes, lasting up to several months and are hard to decompose (Pan et al. 2019, Xu et al. 2021, Zhao et al. 2021).

While studies have reported EPFRs to be pervasive in the atmosphere, little is known about the presence of EPFRs in the household environment. Studies that have explored household environment mostly focused on airborne EPFRs produced from solid fuel combustion used for cooking and heating in China, demonstrating household appliances and behaviours can be important emission sources for residential EPFRs (Jia et al. 2020, Qian et al. 2020, Xu et al. 2020, Zhao et al. 2022, Zhao et al. 2021). However, one study raised interest in other household characteristics such as ventilation (natural ventilation) from air-conditioning and opening of windows in influencing indoor EPFR concentration from the infiltration of ambient TRAP (Sly et al. 2019). Health concerns of EPFR concentration exposure were also raised in that study (Sly et al. 2019), particularly for children due to their increased sensitivity to exposures and continuing respiratory development (Grant et al. 2020). Sly et al. reported that 89 out of 90 house dust samples had detectable EPFRs present, with high EPFR exposure group (≥ 6 × 10^17^ spins/g) associated with a higher number of children reported wheeze compared to low EPFR exposure group (< 4 × 10^17^ spins/g) (Sly et al. 2019). This was the first evidence to link indoor EPFRs in house dust with increased wheeze and supports the suggestion that EPFRs sit on the causal pathway between air pollution and adverse respiratory health outcomes.

The health concerns from EPFR exposure highlights the importance of understanding the factors that influence the presence of EPFRs in indoor environments, particularly in residential homes. We hypothesise that several household characteristics will influence EPFR concentration in household dust. Therefore, this study aimed to understand the concentration of EPFRs in household dust and determine which household characteristics are associated with EPFRs in Australian homes.

## Methods

### Population

The Early Life Lung Function (ELLF) study is a longitudinal birth cohort from Brisbane, Australia that was enrolled during pregnancy and followed until age 7 years. Brisbane is located at the centre of Southeast Queensland and this city has a subtropical climate, with average temperature ranging from 21°C to 30.4°C during summer and 10.5°C to 23.4°C in winter (Bureau of Meteorology 2023). Participants who had previously returned at least one sample of household dust, and who lived within reasonable driving distance from the research centre (around 100km) were invited to participate in home visits following their year 7 follow-up. In total, 50 participants were invited and 24 agreed to participate, one declined, and the rest were non-responders. This home monitoring study was a multiple timepoint (*n* = 2) cross-sectional study, with sampling conducted from February 2021 and concluding in July 2022. A total of 23 participants completed two visits (summer and winter), with only one participant completing a visit in winter and were lost to follow-up for the summer visit. This resulted in a total of 47 individual observations, 23 for summer and 24 for winter.

The Barwon Infant Study (BIS) is a prebirth cohort (n=1074 infants), assembled using an unselected sampling frame, designed to investigate the early life origins of immune dysregulation in the modern environment (Vuillermin et al. 2015). The Barwon region of Victoria, Australia, covers urban, rural, and coastal areas. The Barwon region has a cooler climate on average compared to Brisbane, with temperature ranging from 5.2°C and 14.8°C in winter and 11.9°C to 25.0°C in summer (Bureau of Meteorology 2023). The participant characteristics reflect those of the Australian population overall, although there is a smaller proportion of families from non-English-speaking backgrounds. More details are available in the cohort profile (Vuillermin et al. 2015). Participants were followed-up at multiple time points, including a home visit conducted in a sub-sample (n = 228) at 9 months of age, during which a household assessment was conducted, and a dust sample was collected.

### Air quality

#### Direct measurements

During ELLF home visits, indoor and outdoor air quality were measured simultaneously. Particulate matter (PM) was measured with a TSI DustTrak™ DRX Aerosol monitor 8533 while NO_2_ and CO were measured using a Dräger X-am 5000 with a XXs NO_2_ LC and XXS CO LC sensor. The monitors were set up inside the home in a common area, with a second set placed outside in an area protected from rain. Where possible, sampling occurred over a 24-hour period. Data were collected in 5-minute time averages and averaged across the monitoring period for this study.

#### Modelled air pollution exposure

Direct air quality measurements were not captured in the BIS, so both cohorts were matched to two validated, satellite-based land-use regression models (Sat-LUR) (Knibbs et al. 2014, Knibbs et al. 2018), built for the entire Australian continent from spatial predictors, including land use and satellite information. The residential addresses of each child in both BIS and ELLF were geocoded and matched to an annual estimate of PM_2.5_ and NO_2_ to reflect long-term air pollution exposure (see Supplementary materials).

### Household characteristics survey

The ELLF cohort had previously completed an annual survey, which captured information on housing characteristics (see Table S1). Prior to starting the home visits, we used expert opinion and previous research to determine the household characteristics and behaviours likely to influence indoor air quality, and thus presence of EPFRs. These relationships are depicted in a directed acyclic graph (DAG) (see Figure S1). The ELLF household survey was developed by matching the household survey used in BIS, with a few additional questions included in the ELLF survey. Participants were sent the questionnaire via an online survey system, Qualtrics XM, up to one week prior to each home visit. The responses were checked by a member of the research team and the household characteristics were validated by study staff during each visit. In the BIS, the questionnaire was developed to allow pooling with international cohorts and included validated instruments where possible (Vuillermin et al. 2015). A home visit review was conducted by a trained researcher, including a researcher facilitated survey, and validation of responses by the research team.

### Dust collection

The primary outcome of this study was the presence of EPFRs in household dust. Dust samples were collected from household vacuums by the research team during home visits. Samples were frozen between -22°C to -20°C in the laboratory of Centre for Children’s Health Research, Brisbane or Barwon Biomedical Research Laboratory, Geelong. Samples were shipped to Louisiana State University for analysis. Dust samples were sieved to collect fine dust, and those with weight below minimum sample limit for analysis were excluded. The dust samples underwent electron paramagnetic resonance (i.e., electron spin resonance) to measure EPFR concentration and characteristics. The correction for the content of oxygen-centered radicals (O-EFPRs) in the samples was based on the analysis of the EPFR spectra parameters, using a linear combination of the 3^rd^ power of the g-tensor shift in respect to the position of the pure oxygen-centered radical (2.0049) and spectral broadening represented by the value of delta H peak-to-peak. The resulting adjustment factor was then multiplied by the spin-concentration number. EPFRs and O-EPFRs were reported as the number of spins per gram of dust analysed (spins/g) and analysed as continuous variables. Samples with EPFR concentration below the limit of detection (LOD) were imputed using half the lowest value of EPFR concentration for ELLF (n = 11) and BIS cohort (n = 46) respectively (Hornung & Reed 1990). Samples below the LOD for O-EPFRs were not imputed and excluded from the analysis.

### Statistical analysis

Data were cleaned and summarised using standard descriptive summaries. Household characteristics were categorised depending on the likelihood of generating or circulating air pollutants (see Table S1). There was one record with missing data for cleaning frequency in ELLF and this was imputed with the mean frequency to maintain statistical power. However, a complete case analysis (without imputation) was performed for the BIS cohort due to greater sample size and therefore higher power.

Random forest regression was conducted to determine household characteristics associated with EPFR concentration. Five-fold cross-validation was performed to ensure 100% of the data were being used in building the random forest model. Random forest models with optimal hyper-parameters were analysed for variable importance (see Supplementary materials). Household characteristics that were found to have importance in the random forest model were extracted and Spearman’s rho statistical test was conducted to assess the direction of association. Sensitivity analyses were conducted to determine whether 24-hour ambient PM_2.5_ or annual ambient air pollution (PM_2.5_ and NO_2_) contributed to indoor EPFRs in ELLF cohort. All analyses were conducted in R version 4.1.3 (R Core Team 2022).

## Results

### EPFR in household dust

The EPFRs, air pollution and household characteristics are shown in Table 1 and Table 2. EPFR and O-EPFR concentration were measured and determined in 46 and 35 samples for ELLF cohort; and 136 and 106 samples for BIS cohort respectively. The median (interquartile range, IQR) EPFRs detected in the ELLF households was 1.65 × 10^17^ spins/g (IQR = 3.60 × 10^17^ spins/g) and 5.78 × 10^16^ spins/g (IQR = 1.21 × 10^17^ spins/g) for BIS households (Table 1). Median O-EPFRs measured was 3.41 × 10^16^ spins/g (IQR = 4.70 × 10^16^ spins/g) for BIS and 5.49 × 10^16^ spins/g (IQR = 1.23 × 10^17^ spins/g) for ELLF cohort. Seasonal differences were inconsistent between two geographical cohorts for EPFR concentration. Lower median EPFRs was seen in warmer months of summer (7.06 × 10^14^ spins/g, IQR = 1.83 × 10^17^ spins/g) in comparison to colder months of winter (3.10 × 10^17^ spins/g, IQR = 3.13 × 10^17^ spins/g) for ELLF cohort. BIS cohort demonstrated slightly greater median in summer (6.31 × 10^16^ spins/g; IQR = 4.53 × 10^16^ spins/g) when compared to winter (5.74 × 10^16^, IQR = 1.20 × 10^17^ spins/g). Similar season pattern was observed for O-EPFRs, with greater median concentration in winter for ELLF cohort and in summer for BIS cohort.

**Table 1.** Descriptive results of EPFR concentration, air pollution and season from the Early Life Lung Function (ELLF), Brisbane (n = 46) and Barwon Infant Study (BIS), Geelong, Australia (n = 136).

| Variables | ELLF | BIS |
| --- | --- | --- |
| **<u>Outcomes</u>** |  |  |
| EPFR concentration (spins/g) median (IQR) | $1.65 \times 10^{17}$ ($3.60 \times 10^{17}$) | $5.78 \times 10^{16}$ ($1.21 \times 10^{17}$) |
| EPFR oxygen-weighted concentration (spins/g) median (IQR) | $5.49 \times 10^{16}$ ($1.23 \times 10^{17}$) | $3.41 \times 10^{16}$ ($4.70 \times 10^{16}$) |
| **<u>Air pollution and season</u>** |  |  |
| 24-hour average indoor CO ($\mu\text{g}/\text{m}^3$) mean (SD) | 498.57 (1190.89) | Not collected in BIS |
| 24-hour average indoor NO<sub>2</sub> ($\mu\text{g}/\text{m}^3$) mean (SD) | 4.48 (13.23) | Not collected in BIS |
| 24-hour average indoor PM<sub>2.5</sub> ($\mu\text{g}/\text{m}^3$) mean (SD) | 13.55 (13.19) | Not collected in BIS |
| 24-hour average ambient PM<sub>2.5</sub> ($\mu\text{g}/\text{m}^3$) mean (SD) | 11.63 (6.30) | Not collected in BIS |
| Annual ambient PM<sub>2.5</sub> ($\mu\text{g}/\text{m}^3$) mean (SD) | 6.13 (0.32) | 7.29 (0.73) |
| Annual ambient NO<sub>2</sub> ($\mu\text{g}/\text{m}^3$) mean (SD) | 12.25 (3.97) | 10.13 (2.08) |
| Season of visit n (%) |  |  |
| Summer | 22 (47.8%) | 18 (13.2%) |
| Winter | 24 (52.2%) | 36 (26.5%) |
| Autumn | Not collected in ELLF | 57 (41.9%) |
| Spring | Not collected in ELLF | 25 (18.4%) |

**Table 2.** Descriptive results of household characteristics from the Early Life Lung Function (ELLF), Brisbane (n = 46) and Barwon Infant Study (BIS), Geelong, Australia (n = 136).

| Household characteristics | ELLF | BIS |
| --- | --- | --- |
| **Major house renovation in previous 12 months n (%)** |  |  |
| Yes | 7 (15.2%) | 7 (5.1%) |
| **Type of house n (%)** |  |  |
| Flat/ unit/ apartment | 3 (6.5%) | 2 (1.5%) |
| House | 43 (94.5%) | 134 (98.5%) |
| **House outer wall material n (%)** |  |  |
| Brick | 25 (54.3%) | 93 (68.4%) |
| Weatherboard | 21 (45.7%) | 43 (31.6%) |
| **Type of garage n (%)** |  |  |
| No garage/ carport | 9 (19.6%) | 15 (11.0%) |
| Carport | 7 (15.2%) | 15 (11.0%) |
| Enclosed garage – detached/attached | 30 (65.2%) | 106 (77.9%) |
| **House age mean (SD)** | 43.72 (29.37) | 3.57 (1.24) |
| **Open plan kitchen n (%)** |  |  |
| Yes | 40 (87.0%) | 132 (97.1%) |
| **Fabric flooring in living area n (%)** |  |  |
| Yes (carpet/rug) | 22 (47.8%) | 102 (75.0%) |
| **Presence of indoor fireplace n (%)** |  |  |
| Yes | 8 (17.4%) | 18 (13.2%) |
| **Type of heating in living area n (%)** |  |  |
| Clean heating | 40 (87.0%) | 95 (69.9%) |
| Dirty heating | 6 (13.0%) | 41 (30.1%) |
| **Type of heating in child's room n (%)** |  |  |
| No heating | 24 (52.2%) | 0 |
| Clean heating | 22 (47.8%) | 134 (98.5%) |
| Dirty heating | 0 | 2 (1.5%) |
| **Type of cooling in living area n (%)** |  |  |
| Clean cooling | 28 (60.9%) | 117 (86.0%) |
| Dirty cooling | 18 (39.1%) | 19 (14.0%) |
| **Type of cooling in child's room n (%)** |  |  |
| Clean cooling | 18 (39.1%) | 75 (55.1%) |
| Dirty cooling | 28 (60.9%) | 61 (44.9%) |
| **Type of cook top n (%)** |  |  |
| Electric (ceramic, coil, solid plate) | 32 (69.6%) | 14 (10.3%) |
| Gas | 14 (30.4%) | 122 (89.7%) |
| **Frequency of extractor fan use n (%)** |  |  |
| Always | 19 (41.3%) | 60 (44.1%) |
| Occasionally | 18 (39.1%) | 61 (44.9%) |
| Never | 9 (19.6%) | 15 (11.0%) |
| **Type of oven n (%)** |  |  |
| Electric | 42 (91.3%) | 101 (74.3%) |
| Gas | 4 (8.5%) | 35 (25.7%) |
| **Frequency of candles/incense use at home mean (SD)** | 1.24 (1.49) | 1.32 (1.64) |
| **Mosquito coil use at home n (%)** |  |  |
| Yes | 10 (21.7%) | 5 (3.6%) |
| **Any family members smoke/vape n (%)** |  |  |
| Yes | 4 (8.7%) | 17 (12.5%) |
| **Open windows/door in the past 7 days\* mean (SD)** | 5.76 (2.13) | 2.38 (0.94) |
| **Neighbourhood traffic n (%)** |  |  |
| Little to low | 40 (87.0%) | 77 (56.6%) |
| Some | 3 (6.5%) | 28 (20.6%) |
| High | 3 (6.5%) | 31 (22.8%) |
| **Household size\* mean (SD)** | 4.22 (0.84) | Not collected in BIS |
| **Presence of pets n (%)** |  |  |
| Yes | 36 (78.3%) | Not collected in BIS |
| **Fabric flooring in restroom n (%)** | Not collected in ELLF |  |
| Yes |  | 77 (56.6%) |
| **Fabric flooring in kitchen n (%)** | Not collected in ELLF |  |
| Yes |  | 4 (2.9%) |
| **Fabric flooring in parent's room n (%)** | Not collected in ELLF |  |
| Yes |  | 120 (88.2%) |
| **Fabric flooring in child's room n (%)** | Not collected in ELLF |  |
| Yes |  | 119 (87.5%) |
| **Total cigarette per day mean (SD)** | Not collected in ELLF | 1.56 (4.92) |
| **Method to clean in the house n (%)** |  |  |
| Mop/ vacuum | 39 (84.8%) | 115 (84.6%) |
| Sweep | 7 (15.2%) | 21 (15.4%) |
| **Method to clean surfaces n (%)** |  |  |
| Wet cloth | 25 (54.3%) | Not collected in BIS |
| Dry cloth/ dusting | 21 (45.7%) |  |
| **Frequency of cleaning floors per week (days) mean (SD)** |  |  |
|  | 2.68 (2.91) | Not collected in BIS |
| **Frequency of cleaning surfaces per week (days) mean (SD)** |  |  |
|  | 1.50 (1.53) | Not collected in BIS |
| **Frequency of cleaning floors in living area mean (SD)** | Not collected in ELLF | 3.72 (0.86) |
| **Frequency of cleaning floors in child's room mean (SD)** | Not collected in ELLF | 3.01 (0.76) |

\*ELLF cohort: frequency of candles/incense use at home – 0 = not at all, 1 = every 3-4 months, 2 = 1-2/month, 3 = about 1/week, 4 = a few times/week, 5 = everyday; open windows/door in the past 7 days – number of days windows/ doors opened in the past 7 days; household size – number of family members living at home; BIS cohort: house age – 1 < 2 years, 2 = 2 to 5 years, 3 = 6 to 19 years, 4 = 20 to 49 years, 5 = 50+ years; open windows/door in the past 7 days – 1 = not at all/ < 1 hr/day, 2 = 1-3 hr/day, 3 = 4-12 hr/day, 4 > 12 hr/day; frequency of candles/incense use at home – 0 = not at all, 1 = 1-2 times/month, 2 < 1/week, 3 = about 1/week, 4 = a few times/week, 5 = everyday; frequency of cleaning floors in living area/ child's room – 1 < 1/month, 2 = 1-3 times/month, 3 = 1/week, 4 = a few times/week, 5 = every day.

### Household characteristics associated with EPFR concentration

#### ELLF cohort

This study found half of the (n = 18/36) household characteristics were important contributors to the EPFR concentration in Brisbane’s household dust (see Figure S2). These factors can be categorised into the following themes: house structure, household practices that affect dust resuspension, combustion related activies, ventilation and ambient environmental factors.

Housing structures such as presence of enclosed garage, carport and houses with weatherboard cladding were associated with higher EPFRs; whereas houses without garages and older in age were associated with lower EPFRs (Figure *1*). Units and apartments had lower EPFRs in comparison to detached houses. Factors associated with higher EPFR concentration were household practices that promote dust resuspension (decreased frequency of cleaning and major house renovation) and combustion related activities (lighting candles, incense and occasionally/never using extractor fan). Factors associated with lower EPFR concentration were increased frequency of opening windows/doors, living in a lower traffic neighbourhoods and summer season. Similar results were seen for O-EPFR concentration random forest in ELLF cohort (see Figure S3). Consistent positive associations with O-EPFRs in dust were found for combustion related activities such as dirty heating methods, presence of indoor fireplaces, and use mosquito coils in homes. Higher indoor PM_2.5_ was also associated with higher levels of O-EPFRs. One unusual association observed was lower O-EPFRs in houses with open plan kitchens (Figure 2).

**Figure 1.**
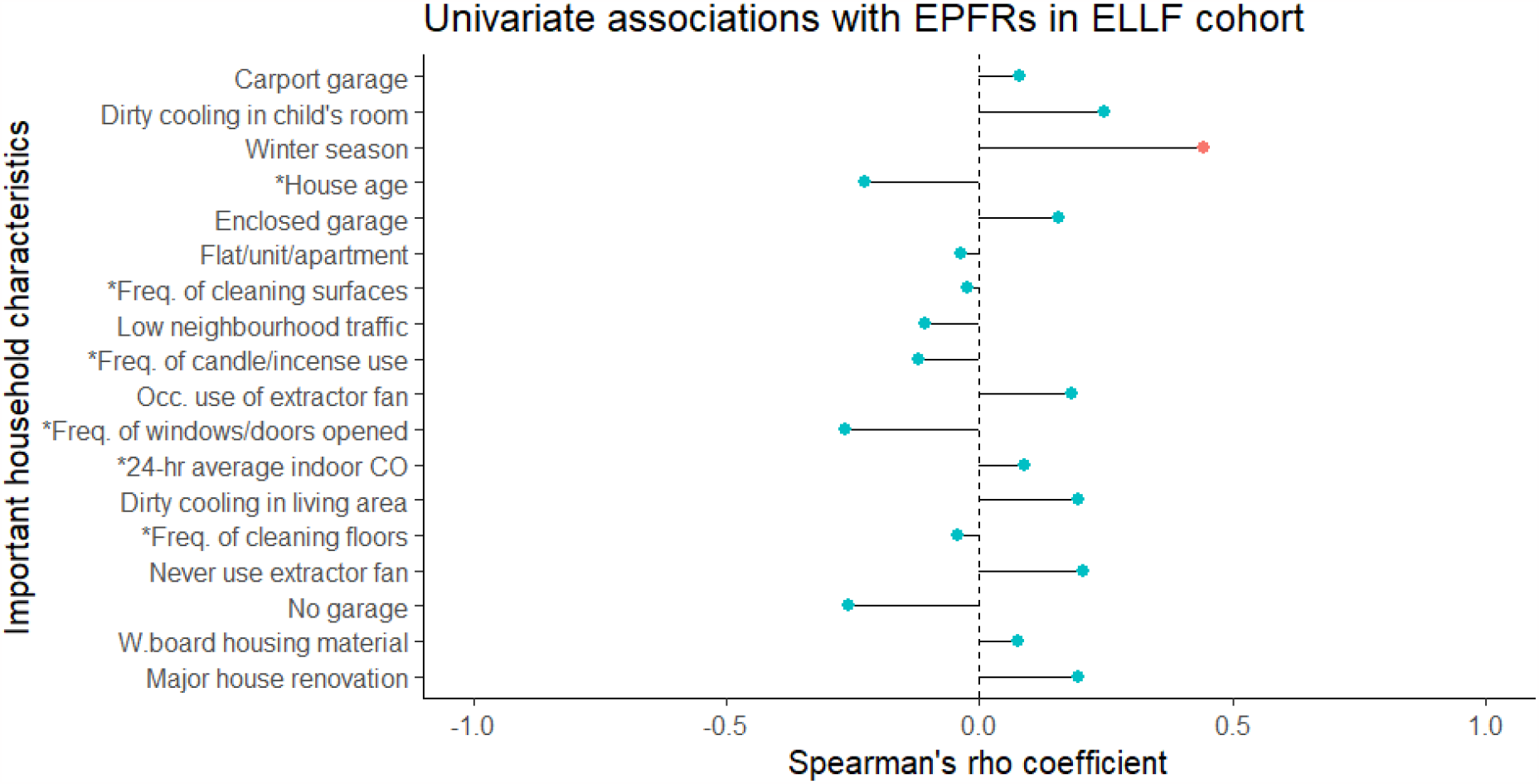
Univariate associations between important household characteristics with EPFR concentration identified from random forest model in ELLF cohort (*indicates continuous variable. The rest are binary rank variable. Red point signifies statistically significant association).

**Figure 2.**
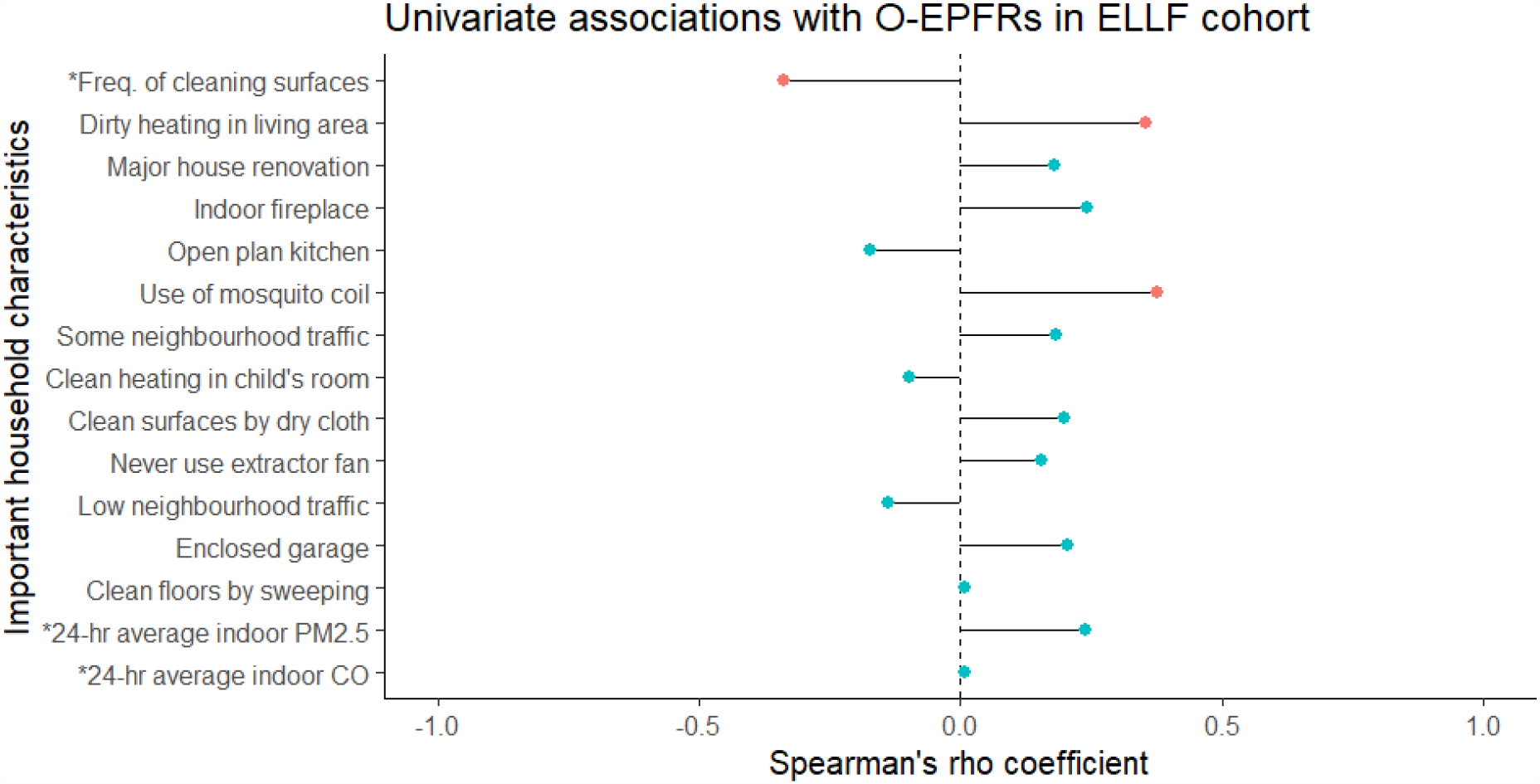
Univariate associations between important household characteristics with EPFR oxygen-weighted concentration identified from random forest model in ELLF cohort (* indicates continuous variable. The rest are binary rank variables. Red point signifies statistically significant association).

**Figure 3.**
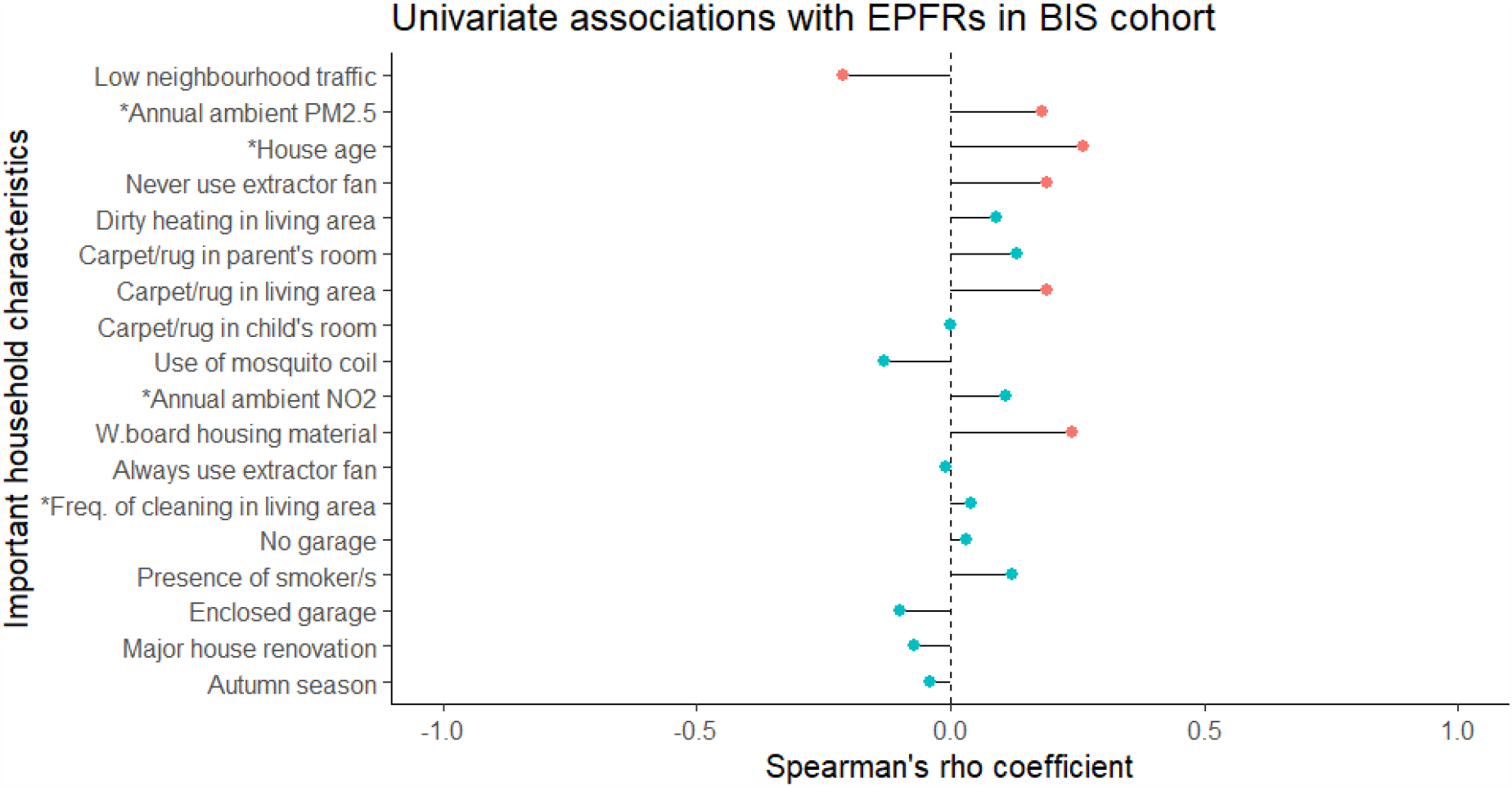
Univariate associations between important household characteristics with EPFR concentration identified from random forest model in BIS cohort (*indicates continuous variable. The rest are binary rank variable. Red point signifies statistically significant association).

### BIS cohort

In the BIS cohort, the regression random forest found that the most important variables represented for EPFR concentration was TRAP (Figure *3*). Higher EPFR concentration was associated with higher neighbourhood traffic and ambient air pollution (PM_2.5_ and NO_2_). Further, house age, never using extractor fan when cooking, increased combustion related activities (presence of smoker/s in home and dirty heating method used in living area) and presence of fabric flooring (carpet/rug in parent’s room and living room) were also associated with higher EPFR concentration in house dust. Lower EPFR concentration was observed in autumn. A few unexpected directions of associations were observed such as houses without attached garages and higher cleaning frequency were seen to slightly increase EPFR concentration, while houses that recently undergone major house renovation, have enclosed garages and use of mosquito coils were associated with lower EPFRs. Household appliances such as type of cook top and oven were not associated with EPFRs nor O-EPFRs in household dust (see Figure S4 and Figure S5).

As with EPFR concentration, TRAP was the most important characteristic related to O-EPFRs (Figure *4*). Higher O-EPFRs were associated with older homes, carports, carpets or rugs in kitchens and parent’s bedrooms, number of cigarettes per day, dirty heating method in the living rooms and use of gas ovens. Lower O-EPFRs were associated with major renovations, carpets or rugs in child’s bedrooms and winter season.

**Figure 4.**
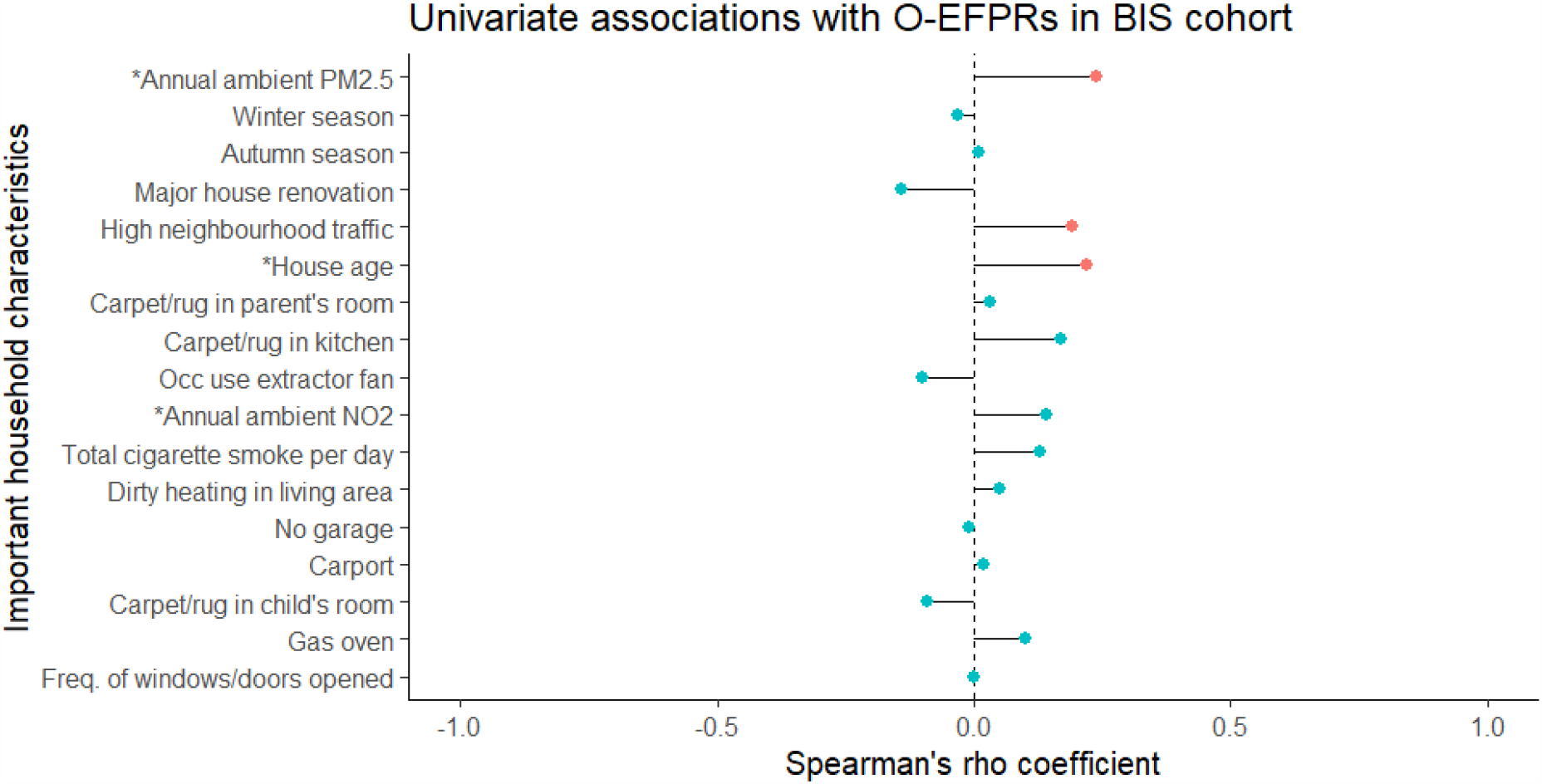
Univariate associations between important household characteristics with EPFR oxygen-weighted concentration identified from random forest model in BIS cohort (*indicates continuous variable. The rest are binary rank variable. Red point signifies statistically significant association).

### Comparison between ELLF and BIS cohorts

We compared the household characteristics associated with EPFRs and O-EPFRs in both cohorts to assess their similarity. Eight household characteristics in the ELLF households shared the same EPFR concentration importance in BIS (Table 3). Only two household characteristics shared the same O-EPFR concentration importance between both cohorts. Among all the household characteristics, recent major renovation was the most consistent variable, showing importance in both cohorts and with both EPFRs concentration and O-EPFRs

**Table 3.**
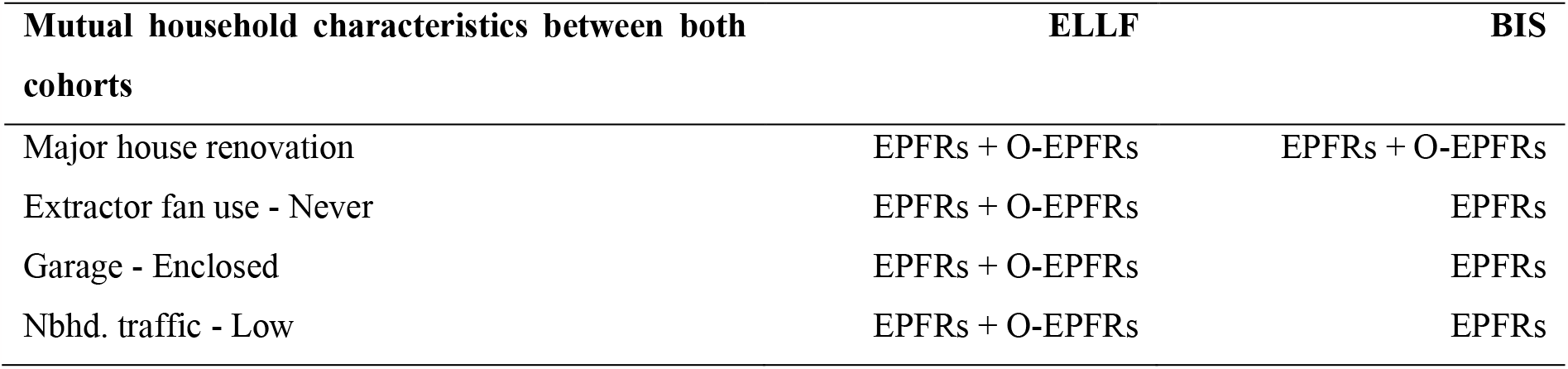
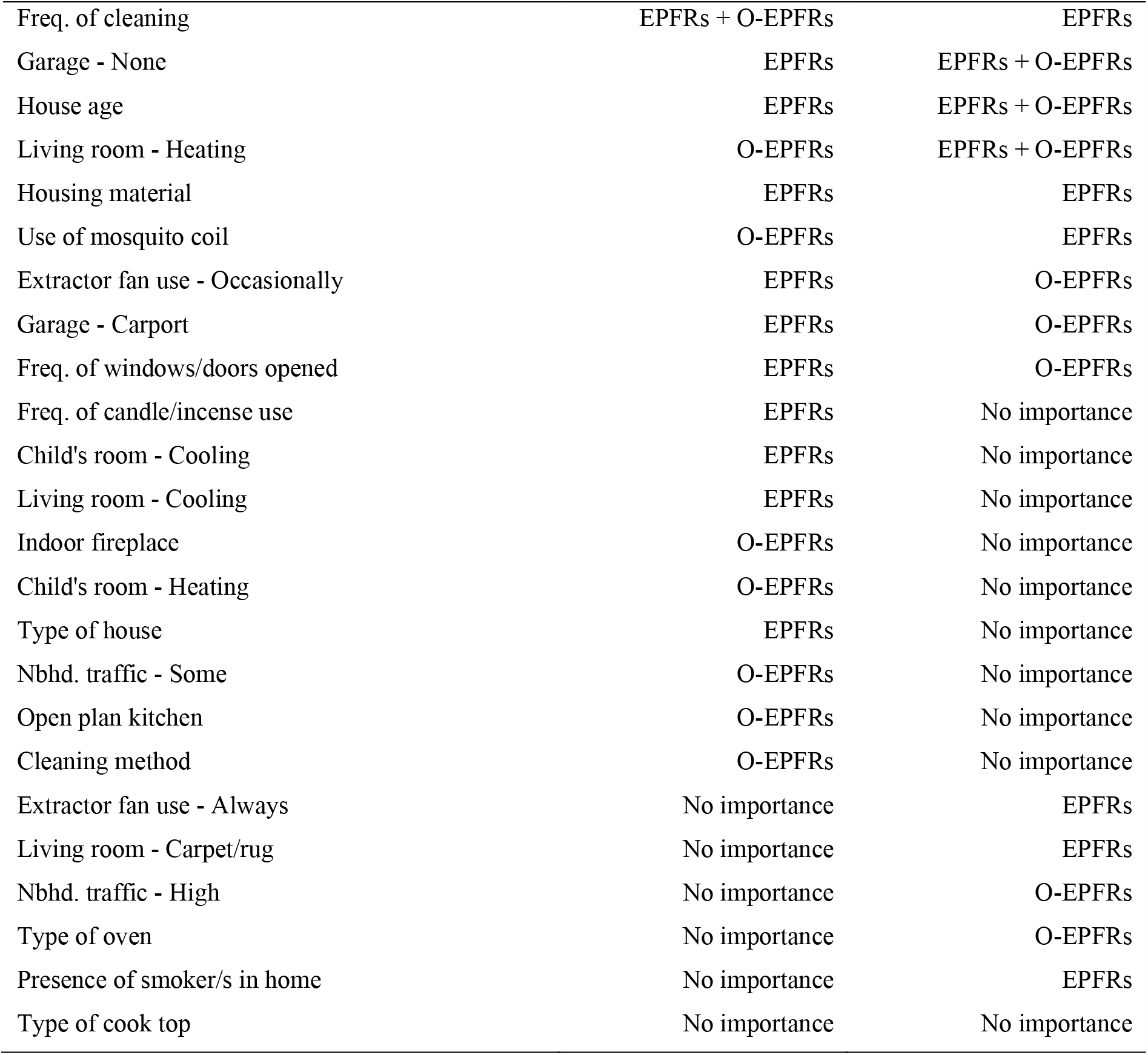
Household characteristics found important across all random forest models in ELLF and BIS cohort.

Important household characteristics (as identified in random forest model) for both cohorts were major house renovation, never using extractor fan when cooking, presence of enclosed garage, low neighbourhood traffic, frequency of cleaning, no garage, age of house, housing material and heating method used in living area.

However, correlation assessment showed consistent direction only for homes built with weatherboard cladding, households never using extractor fan when cooking and low neighbourhood traffic for EPFR concentration (Figure 5), and heating method used in living area for O-EPFRs (Figure 6).

**Figure 5.**
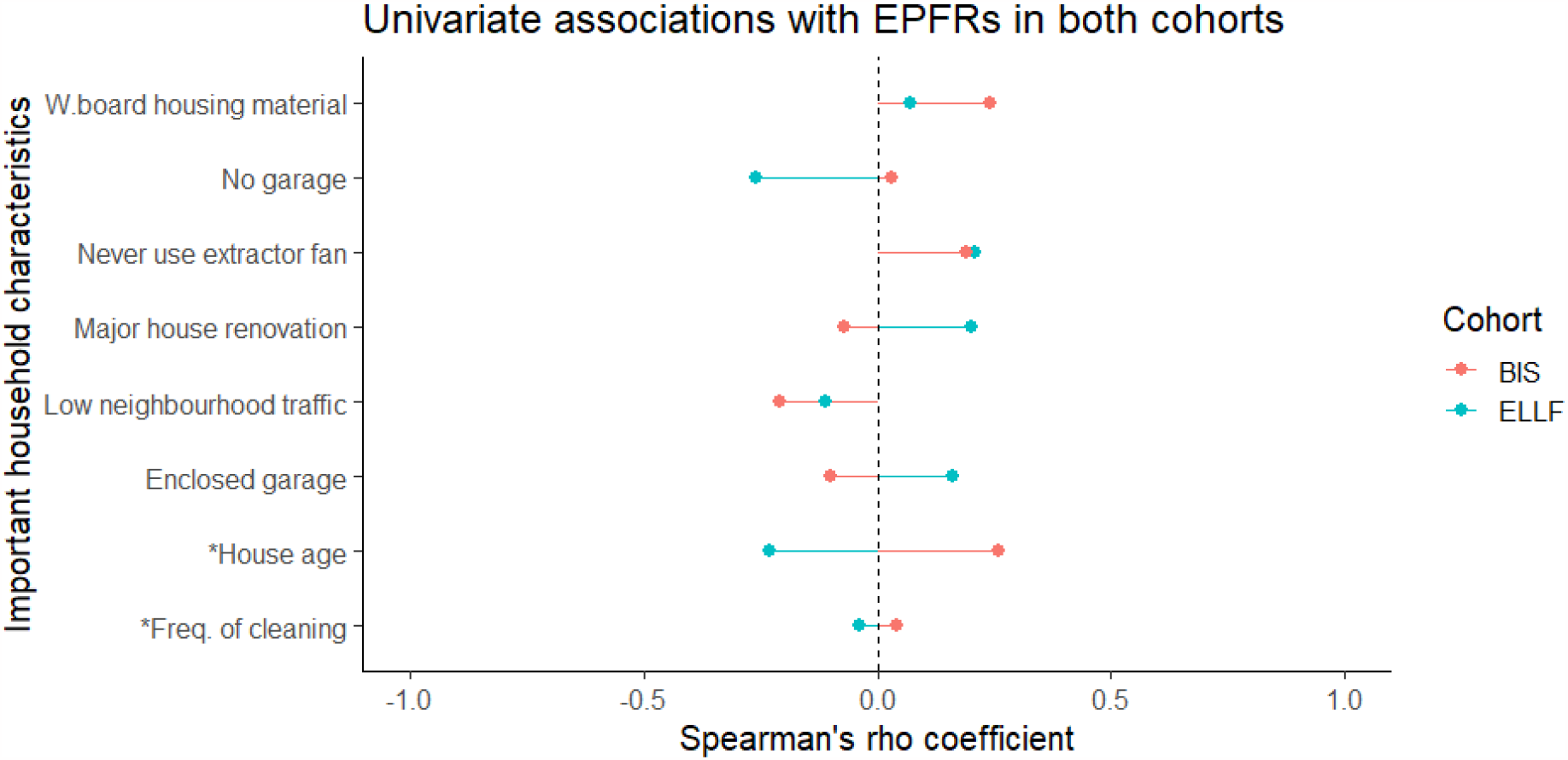
Univariate associations between important household characteristics with EPFR concentration identified from ELLF and BIS random forest models (*indicates continuous variable).

**Figure 6.**
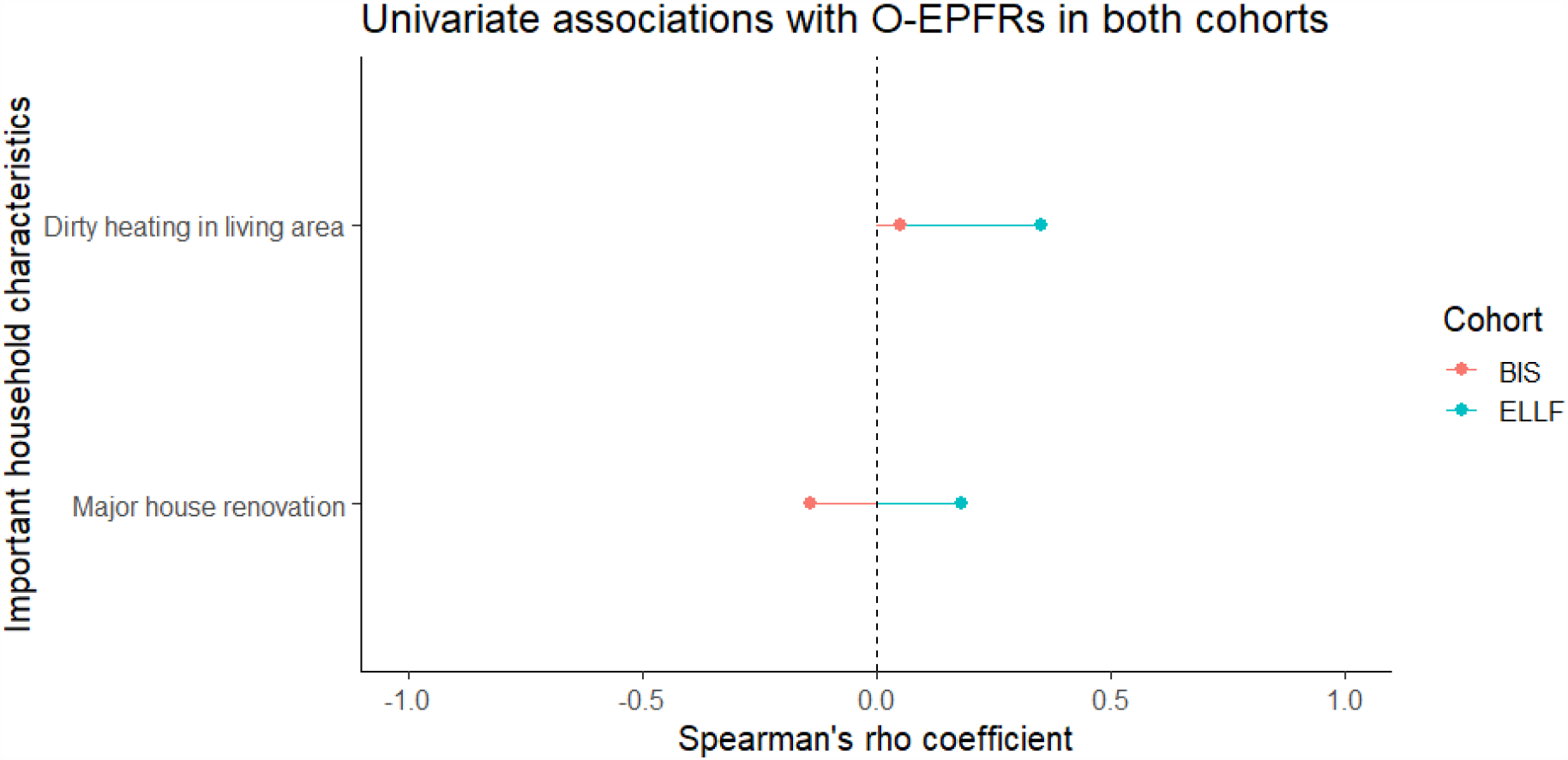
Univariate associations between important household characteristics with EPFR oxygen-weighted concentration identified from ELLF and BIS random forest models.

### Sensitivity analysis

We explored the effect of modelled vs measured ambient PM_2.5_ in the ELLF cohort. Neither variable was important in the analyses of EPFRs and O-EPFRs (see Supplementary materials). Modelled ambient NO_2_ in ELLF cohort demonstrated importance in O-EPFRs but little importance in EPFRs.

## Discussion

The discovery of environmentally persistent free radicals is a relatively new phenomenon, and research exploring indoor sources of EPFRs concentration are in its infancy. To the best of our knowledge, the present study is the first study to explore household characteristics associated with EPFRs and O-EPFRs in household dust. We found that housing structures, TRAP, heating method, house ventilation and household activities that affect dust circulation were the most important household characteristics associated with EPFRs in Australian household dust.

EPFRs are bounded to PM and a large body of literature has established vehicle emissions as one of the major sources of EPFRs on airborne particles (Chen et al. 2019, Wang et al. 2020). A study which investigated EPFR concentrations in different locations found that EPFR signals were less prominent in agricultural areas compared to residential areas due to lower vehicle exhaust emissions (Wang et al. 2020). The results of the present study support these findings, with lower EPFRs in both ELLF and BIS cohorts for those households located on streets with low neighbourhood traffic. In the BIS cohort, annual ambient NO_2_ and PM_2.5_ were associated with EPFRs in dust. However, only ambient NO_2_, was associated with EPFRs in ELLF homes.

In the present study, the physical structures of a house, such as type of garage, age of house and housing material were associated with EPFRs in household dust. It is likely that the physical structures of a home influence the overall air tightness (Ambrose & Syme 2015, Mallach et al. 2017, Oh & Kim 2020). The air exchange rate (ACH) is typically used to understand the leakiness of a house by determining the rate of outdoor air replacing indoor air in a specified space (Guo et al. 2008). Two Australian reports have found older houses in Australia tended to have higher ACH, signifying higher outdoor air penetration than newer houses (Ambrose & Syme 2015, Metropolitan Fire and Emergency Services Board 2011). Further, houses built with weatherboard cladding have higher ACH values than brick houses (Metropolitan Fire and Emergency Services Board 2011). Higher air penetration for houses constructed from timber may be due higher likelihood of termite damages, sunlight, and water damage (Reardon et al. 2020). We expected to observe higher EPFRs for houses older in age and built with weatherboard due to greater ambient air penetration and our results were consistent with our hypothesis. However, the findings between age of house and EPFR concentration were inconsistent between the cohorts, suggesting the construction materials and design may be more important than age. The type of garage can influence indoor air quality. It has previously been shown that attached garages or underground parking garages led to higher concentration of pollutants (volatile organic compounds, PM and CO) inside the houses when compared to non-attached garages (Mallach et al. 2017, Oh & Kim 2020). Pollutants generated in enclosed and attached garages from the car can accumulate and penetrate inside the home through walls and ceilings (Mallach et al. 2017, Oh & Kim 2020). We found an association between enclosed garages and higher EPFR concentration in the ELLF cohort, but not in the BIS study.

The heating method used in homes was an important factor in EPFR concentration, particularly for the more redox-active O-EPFRs. Current studies conducted in China found that EPFRs were greater in colder months, with a study estimating an equivalent EPFR exposure found in 23-73 cigarettes per day (Chen et al. 2019, Jia et al. 2023, Xu et al. 2021, Xu et al. 2020, Zhao et al. 2022). The authors suggested that the high central heating use such as coal combustion to keep residential homes warm during winter can explain the higher EPFR concentration as combustion of coals and fuels are an established source of EPFR. In addition, studies have identified that EPFR concentration with an adjacent oxygen atom was more prominent in colder weather, suggesting incomplete combustion (Jia et al. 2023, Xu et al. 2021, Zhao et al. 2022). Consistent with the existing literature, our results supported their hypothesis, demonstrating dirty type of heating method used such as gas heater, oil/kerosene/diesel heater and fireplace to be positively and significantly associated with O-EPFRs in both cohorts. However, seasonal difference in our results were inconclusive with only Brisbane displaying greater EPFR concentration in winter than summer. In addition, cigarette smoking is an established combustion activity that generate EPFRs (Gehling & Dellinger 2013, Xu et al. 2021), but our analyses demonstrated that smoking households was only found to be important in BIS cohort. The potential significance of cigarette smoking was not adequately assessed in ELLF, possibly due to different level of exposure as family members reported smoking outdoors in the ELLF households.

Our results also discovered that use of extractor fan when cooking was an important household characteristic associated with EPFRs. Extensive research on cooking fumes by-products have found that toxic compounds such as polycyclic aromatic hydrocarbons (PAH), aldehydes, alkanoic acids, heterocyclic aromatic amine and other harmful products can be released when cooking foods in higher temperature (Chiang et al. 1999, Mehta 2015, Svedahl et al. 2009). It is established in the literature that EPFR concentration can be generated from solid fuel combustion used for cooking (Zhao et al. 2022, Zhao et al. 2021), however, there is other research that found a close co-existence of EPFRs and PAHs (Jia et al. 2018, Lammel et al. 2020, Sun et al. 2022). Regardless of whether EPFR was generated from the fuel source used in cooking or from heating the food in high temperature, EPFR produced from cooking is evident, and the use of extractor fan can act to reduce the association between cooking EPFRs and its concentration in the house. In our study, we found that Australian homes that never use their extractor fans when cooking had higher EPFR concentration. We hypothesise that EPFRs generated from cooking can be lowered in the house when extractor fans are used regularly as previous studies looking at similar concept of extractor fumes exhibited reduction of harmful compounds and lower risk of lung cancer when fume extraction was used during cooking (Chiang et al. 1998, Ko et al. 1997).

Lastly, activities affecting dust resuspension in the house were found to be important household characteristics of EPFRs in Australian household. Effective cleaning habits can reduce dust concentration and indoor air pollutants (Roberts et al. 2009), and we hypothesise that this can potentially lower EPFRs generated from indoor combustion or infiltration of outdoor air pollution. However, our results showed conflicting results with only ELLF cohort demonstrating higher EPFRs for houses that do not practice regular cleaning. Research on increased small PM (aerodynamic diameter of 1.7 μm) from houses undergone construction (Abdel Hameed et al. 2004) and EPFRs has not been extensively studied. Current knowledge of EPFRs production is generated through combustion (Gehling & Dellinger 2013, Sly et al. 2019, Vejerano et al. 2018, Xu et al. 2021, Zhao et al. 2021), therefore, more research is warranted to understand the importance and causal pathway of increased PM from major construction and EPFR concentration.

While results demonstrated that indoor EPFRs can be generated from household activities, TRAP components also have importance in influencing EPFRs concentration within households. Our research provided some insight on the potential of household characteristics that may affect EPFR concentration, which can potentially cause adverse health impacts. Future research should link our research findings of EPFRs influenced by housing structures, TRAP, heating method, house ventilation and household activities affecting dust resuspension to health effects.

There are strengths and limitations to this study. Firstly, we anticipated that the small number of participants recruited in ELLF cohort would be a potential issue in our analysis; therefore, 1) we validated our data with a larger cohort (BIS cohort) in Geelong and 2) we conducted an ensemble machine learning method known as random forest model to allow us to gain good statistical insight from our highly dimensioned data. While this method can handle small sample sizes with large number of predictors, there is need to conduct a larger study to establish the findings of this research. In addition, this study was a prospective study which eliminated any concerns on recall biases and can provide a more valid information on exposures. Furthermore, the air pollution measured in participants’ homes for ELLF cohort were collected using our own monitors. The use of our own equipment in each house provides a better reflection of personal pollutant exposures when compared to data collected from public databases.

## Conclusion

This study found that house age, type of garages, building material, heating methods, extractor fan practices when cooking, TRAP, recent major house renovation and cleaning habits are important household characteristics associated with EPFRs in Australian homes. Some of these household characteristics are behaviours and activities that can be modified to lower the presence of EPFRs inside a home.

## Data Availability

Data are only available after approval by the study chief investigator team

## Acknowledgements

The author would like to thank Dr Trish Gilholm for her expertise, time and advice on the modelling of our random forests.

## Statements & Declarations

### Funding

Data collection was supported by the National Institute of Environmental Health Sciences of the National Institutes of Health under Award Number P42ES013648. The content is solely the responsibility of the authors and does not necessarily represent the official views of the National Institutes of Health.

### Competing interests

The authors have declared that there was no conflict of interests that may have influenced the work in this paper.

### Author contributions

Dwan Vilcins, Peter D Sly, Stephania Cormier and Slawo Lomnicki contributed to the study conception and design. Material preparation, data collection and analysis were performed by Dwan Vilcins and Wen Ray Lee. Prakash Dangal processed the dust samples and measured the presence of EPFRs. Manuscript draft was written by Wen Ray Lee and all authors have commented and approved of the final manuscript.

## Notes

### Competing Interest Statement

The authors have declared no competing interest.

### Author Declarations

The Childrens Health Queensland Human Research Ethics Committee reviewed and gave ethical approval for the study (HREC/13/QRCH/156).

