## Supplementary materials for "Household characteristics associated with environmentally persistent free radicals in house dust in two Australian locations"

**Methods**

**Directed Acyclic Graph (DAG)**

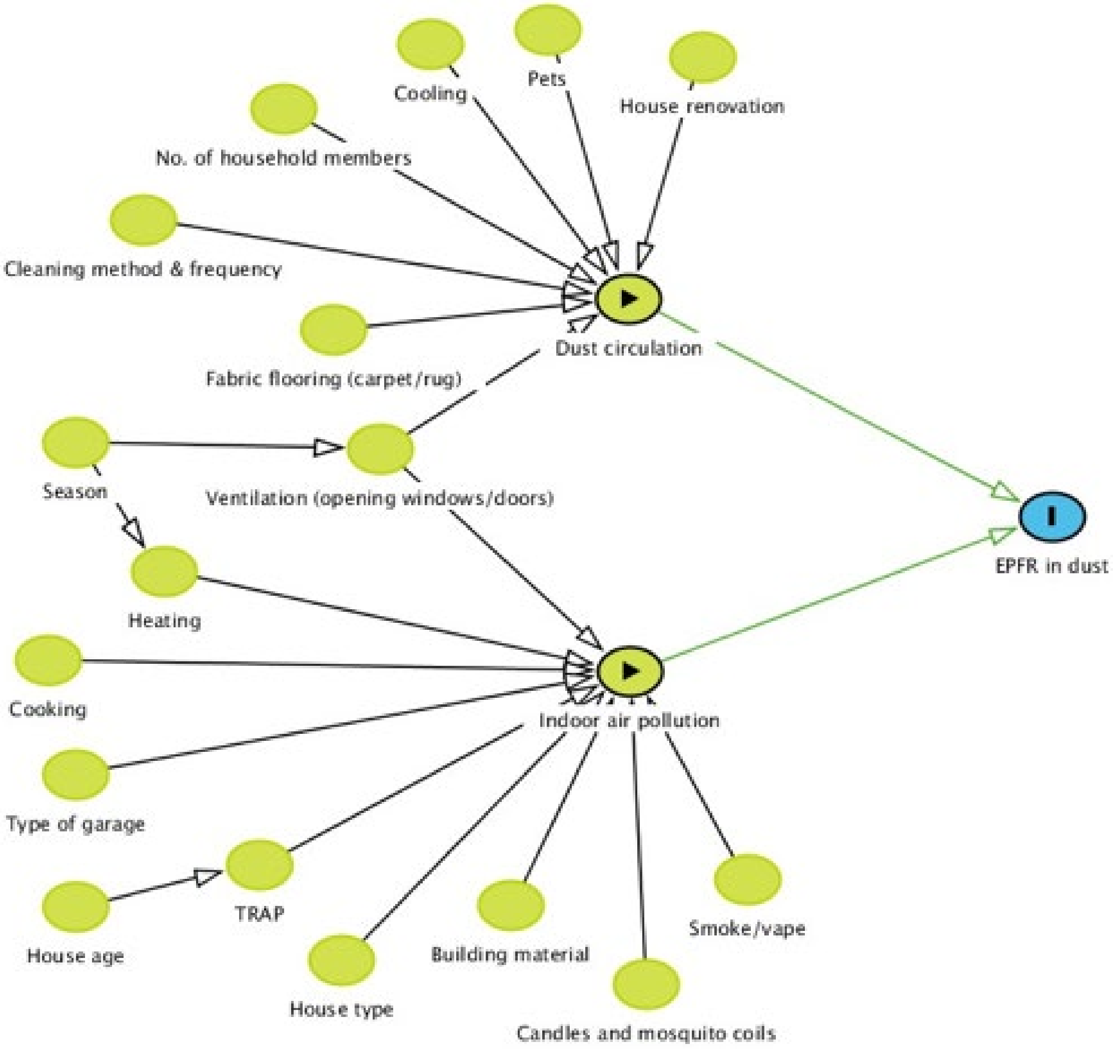


Figure S1 Directed Acyclic Graph of household characteristics and presence of EPFR in dust.

**Direct measurements of air pollution exposure**

During ELLF home visits, particulate matter (PM) was measured with a TSI DustTrak$^{TM}$ DRX Aerosol monitor 8533 which uses a light-scattering laser photometer to simultaneously measure size and mass of airborne particles.

**Modelled air pollution exposure**

The Sat-LURs provide an annual mean concentration of $PM_{2.5}$ and $NO_2$ with a spatial resolution ranging from 100 m in urban areas to 500 m in rural/remote areas. The $NO_2$ model captured between 66% and 81% of variability in measured annual $NO_2$ concentrations, respectively, with RMSE of 1.4 to 2 ppb over the time span of the cohort. The $PM_{2.5}$ model captured between 52% and 63% of variability in $PM_{2.5}$, with an RMSE of 1 to 1.2 $\mu g/m^3$ (Knibbs et al. 2018). The residential addresses of each child in both BIS and ELLF were geocoded to 6 decimal places and matched to an annual estimate of $PM_{2.5}$ and $NO_2$ to reflect long-term air pollution exposure.

**Statistical analysis**

Random forest parameters such as number of trees, minimum number of observations in a terminal node, maximum number of terminal nodes and number of variables at each split were determined based on the best mean RMSE (Root-mean-square error) using grid search from a custom algorithm built in '*caret*' package. Random forest models with optimal hyper-parameters were analysed for variable importance. The assessment of *%IncMSE* (Increase of Mean Square Error) is evaluated based on how much poorer the prediction and performance of the model if the data of the respective predictor were randomly permuted. The poorer the performance of the model, indicated by greater *%IncMSE,* the more important the household variable in determining indoor EPFRs. Only household variables with *%IncMSE* greater than 0.05 were deemed important in this paper as percentage below the latter value signifies negligible importance.

**Household characteristics survey**

Table S1 Household characteristics determined from questionnaire responses conducted in ELLF and BIS cohorts.

| | | Survey question description | |
|---|---|---|---|
| **Variables** | **Category** | **ELLF questionnaire** | **BIS questionnaire** |
| Major house renovation | Yes | Q: Any major renovation involving construction in the past 12 months?<br>R: “Main living room”; “Child’s bedroom”; “Kitchen” | Q: Have any of the following rooms (baby’s room/your bedroom/your living room/your kitchen) in your home been decorated since your baby’s last review at 6 months of age?<br>R: “Yes” and “Major renovation (involving construction)” |
| Type of house | Flat/unit/apartment | Q: What type of home do you have?<br>R: “Unit/ apartment or flat” | Q: What kind of building is your home?<br>R: “House – Detached/Semi-detached/Terrace” |
| | House | Q: What type of home do you have?<br>R: “Detached”; “Semi-detached”; “Terrace” | Q: What kind of building is your home?<br>R: “Flat unit or apartment” |
| House outer wall material | Brick | Q: What is the building material of the outer walls?<br>R: “Brick” | Q: Building material of the outer walls<br>R: “Brick”; “Other (“Brick veneer”, “Stone brick”) |
| | Weatherboard | Q: What is the building material of the outer walls?<br>R: “Weatherboard”; “Other” (“Concrete block on lower storey and weather board on second storey”) | Q: Building material of the outer walls<br>R: “Weatherboard”; “Fibro”; “Other (“hardie plank”; “corrugated iron”; “cladding”; “timber”) |
| Type of garage | No garage/carport | Q: What type of garage or carport do you have?<br>R: “No garage or carport” | Q: Do you have a garage or carport?<br>R: “No garage or carport” |
| | Carport | Q: What type of garage or carport do you have?<br>R: “Carport” | Q: Do you have a garage or carport?<br>R: “Carport” |
| | Enclosed garage – detached/attached | Q: What type of garage or carport do you have?<br>R: “Enclosed garage, detached”; “Enclosed garage, attached” | Q: Do you have a garage or carport?<br>R: “Enclosed garage – attached”; “Enclosed garage, detached”; “Enclosed garage – but not used to park motor” |
| House age | Continuous | Q: How old is your house (in years)? If you are not sure, please provide an estimate age | Q: How old is your hoe (approximately)? Please consider when the building was first constructed, not when it was remodelled, added or converted/<br>R: “Less than 2 years = 1”; “2-5 years = 2”; “6-19 years = 3”; “20-49 years = 4”; “50 years or more = 5” |
| Open plan kitchen | Yes | Q: Do you have an open-plan kitchen (i.e. attached to the living quarters)<br>R: “Yes” | Q: Is the kitchen open-plan (i.e. open to the main living area)?<br>R: “Yes” |
| Fabric flooring in living area | No | Q: What type of flooring?<br>R: “Tiles/slate/cork”; “Linoleum”; “Wooden floor”; “Concrete floor” | Q: What kind of floor coverings do you have indoors in your living room?<br>R: “Tile”; “Wood”; “Linoleum”; “Concrete”; “Slate” |
| | Yes | Q: What type of flooring?<br>R: “Carpet; Rug” | Q: What kind of floor coverings do you have indoors in your living room?<br>R1: “Carpet”; “Rug”<br>R2: Have both fabric and non-fabric flooring responses |
| | Yes | Q: Do you have an indoor fireplace?<br>R: “Yes” | Q: What type of heating do you us in your main living area *(e.g. living room)?<br>R: “Open fireplace”; “Closed fireplace” |

| | | | |
|---|---|---|---|
| Type of heating in living area | Clean heating | Q: What type of heating do you use in your main living area?<br>R: “No heating”; “Central heating (ducted air)”; “Central heating (slab/underfoot)”; “Reverse-cycle air-conditioning”; “Electric heater” | Q: What type of heating do you us in your main living area *(e.g. living room)? *main source of heating was chosen*<br>R: “Central heating (ducted air)”; Central heating (in slab/underfloor)”; “Central heating (hydronic/hot water)”; “Reverse-cycle air-conditioning unit”; “Electric convection/column heater”; “Electric panel heater”; “Electric fan heater”; “Electric bar heater”; “Other (“Concrete slab heating”) |
| | Dirty heating | Q: What type of heating do you use in your main living area?<br>R: “Gas heater with a flue”; “Gas heater without a flue”; “Oil/kerosene/disesel heater”; “Open fireplace”; ‘Closed fireplace” | Q: What type of heating do you us in your main living area *(e.g. living room)? *main source of heating was chosen*<br>R: “Gas heater with a flue”; “Gas heater without a flue”; “Oil/kerosene/diesel heater”; “Open fireplace”; “Closed fireplace” |
| Type of heating in child’s room | No heating | Q: What type of heating do you use in your child’s bedroom?<br>R: “No heating” | *No household with no heating in baby’s room* |
| | Clean heating | Q: What type of heating do you use in your child’s bedroom?<br>R: “Central heating (ducted air)”; “Central heating (slab/underfoot)”; “Reverse-cycle air-conditioning”; “Electric heater” | Q: When you heat your baby’s bedroom at night, what types of heating do you use? *main source of heating was chosen*<br>R: “Central heating (ducted air)”; Central heating (in slab/underfloor)”; “Central heating (hydronic/hot water)”; “Reverse-cycle air-conditioning unit”; “Electric convection/column heater”; “Electric panel heater”; “Electric fan heater”; “Electric bar heater”; “Other – Ceramic wall tile” |
| | Dirty heating | *No household with dirty heating in child’s room* | Q: When you heat your baby’s bedroom at night, what types of heating do you use? *main source of heating was chosen*<br>R: “Gas heater with a flue”; “Gas heater without a flue”; “Oil/kerosene/diesel heater”; “Open fireplace”; “Closed fireplace” |
| Type of cooling in living area | Clean cooling | Q: What type of cooling do you use in your main living area?<br>R: “No cooling”; “Central cooling (ducted air)”; “Evaporative air-conditioning”; “Refrigerated air-conditioning”: “Reverse-cycle air-conditioning” | Q: What type of cooling do you use in your main living area? **Mark all that apply.**<br>R: “Ducted central air-conditioning”; “Fixed wall air-conditioning unit”; “Fixed window air-conditioning unit”; “Portable air-conditioning unit”; “No cooling” |
| | Dirty cooling | Q: What type of cooling do you use in your main living area?<br>R1: “Free standing air cooler”; “Ceiling fan”; “Portable/pedestal fan”<br>R2: If both clean and dirty cooling methods are used | Q: What type of cooling do you use in your main living area? **Mark all that apply.**<br>R1: “Ceiling fan”; “Portable fan”<br>R2: If both clean and dirty cooling methods are used |
| Type of cooling in child’s room | Clean cooling | Q: What type of cooling do you use in your child’s bedroom?<br>R: “No cooling”; “Central cooling (ducted air)”; “Evaporative air-conditioning”; “Refrigerated air-conditioning”: “Reverse-cycle air-conditioning” | Q: When you cool your baby’s bedroom at night, what types of cooling do you use? **Mark all that apply.**<br>R: “Ducted central air-conditioning”; “Fixed wall air-conditioning unit”; “Fixed window air-conditioning unit”; “Portable air-conditioning unit”; “No cooling” |
| | Dirty cooling | Q: What type of cooling do you use in your child’s bedroom?<br>R1: “Free standing air cooler”; “Ceiling fan”; “Portable/pedestal fan”<br>R2: If cleaning and dirty cooling are used | Q: When you cool your baby’s bedroom at night, what types of cooling do you use? **Mark all that apply.**<br>R1: “Ceiling fan”; “Portable fan”<br>R2: If both clean and dirty cooling methods are used |
| Type of cook top | Electric | Q: What type of cook top do you have at your current residence?<br>R: “Electric (ceramic, coil, solid plate)”; “Induction” | Q: What type of cook top do you have? **Mark all that apply.**<br>R: “Electric”; “Other (“Induction”)” |

| | | | |
|---|---|---|---|
| | Gas | Q: What type of cook top do you have at your current residence?<br>R: "Gas" | Q: What type of cook top do you have? **Mark all that apply.**<br>R: "Gas" |
| Frequency of extractor fan use | Always | Q: How often do you use your extractor hood when cooking on the stovetop?<br>R: "Every time" | Q: How often do you use extractor fan/extractor hood when cooking?<br>R: "Every time" |
| | Occasionally | Q: How often do you use your extractor hood when cooking on the stovetop?<br>R: "Sometimes"; "Usually (half the time or more)" | Q: How often do you use extractor fan/extractor hood when cooking?<br>R: "Sometimes (less than half the time)"; "Usually (half the time or more)" |
| | Never | Q: How often do you use your extractor hood when cooking on the stovetop?<br>R: "Never" | Q: How often do you use extractor fan/extractor hood when cooking?<br>R: "Never" |
| Type of oven | Electric | Q: What type of oven do you have at your current residence?<br>R: "Electric" | Q: What type of oven do you have? **Mark all that apply.**<br>R: "Electric"; "Other ("Forced fan")" |
| | Gas | Q: What type of oven do you have at your current residence?<br>R: "Gas" | Q: What type of oven do you have? **Mark all that apply.**<br>R1: "Gas"<br>R2: Have both electric and gas oven. |
| Frequency of candles/ incense use at home | Continuous | Q: How often have you used candles/incense in your home, in the last 12 months?<br>R: "Everyday = 5"; "A few times a week = 4"; "About once a week = 3"; "1-2 times a month = 2"; "Every 3-4 months = 1"; "Not at all = 0" | Q: Since your baby's 6 month review, how often you used candles or incense in your home?<br>R: "Every day = 5"; "A few times a week = 4"; "About once a week = 3"; "Less than once a week = 2"; "1-3 times a month = 1"; "Not at all = 0" |
| Mosquito coil use at home | No | Q: How often have you used mosquito coil in your home, in the last 12 months?<br>R: "Not at all" | Q: Since your baby's 6 month review, how often you used mosquito coil in your home?<br>R: "Not at all" |
| | Yes | Q: How often have you used candles/incense in your home, in the last 12 months?<br>R: "Everyday"; "A few times a week"; "About once a week"; '1-2 times a month"; "Every 3-4 months" | Q: Since your baby's 6 month review, how often you used mosquito coil in your home?<br>R: "Every day"; "Less than once a week"; "About once a week"; "Less than once a week"; "1-3 times a month" |
| Any family members smoke/vape | Yes | Q: "Does any member of the household smoke or vape?"<br>R: "Yes" | Q: Do you and your partner currently smoke cigarettes and/or any tobacco products?<br>R: "Yes" |
| Open windows/door in the past 7 days | Continuous | Q: In the last 7 days, how often did you open your windows or doors for ventilation?<br>R: "None = 0"; "1 Day = 1"; "2 Days = 2"; "3 Days = 3"; "4 Days = 4"; "5 Days = 5"; "6 Days = 6"; "Everyday = 7 Days = 7" | Q: Thinking about the last 7 days, approximately how many hours a day did you keep any external windows or doors open in your home (for ventilation or let air in)?<br>R: "Not at all/ Less than 1 hour per day = 1"; "1-3 hours per day = 2"; "4-12 hours per day = 3"; "More than 12 hours per day = 4" |
| Neighbourhood traffic | Little to low | Q: What type of street is your home located on?<br>R: "Neighbourhood or local street" | Q: There is heaving traffic on my street or road.<br>R: "Strongly disagree"; "Disagree" |
| | Some | Q: What type of street is your home located on?<br>R: "District road" | Q: There is heaving traffic on my street or road.<br>R: "Neither agree nor disagree" |
| | High | Q: What type of street is your home located on?<br>R: "Suburban road" | Q: There is heaving traffic on my street or road.<br>R: "Strongly agree"; "Agree" |
| Household size | Continuous | Q: How many adults and children usually live in the house? | *Not collected in BIS* |

| | | | |
|---|---|---|---|
| Presence of pets | No | Q: Do you have any pets? (Please include all pets that have lived in your household in the last 12 months)<br>R: “No” | *Not collected in BIS* |
| | Yes | Q: Do you have any pets? (Please include all pets that have lived in your household in the last 12 months)<br>R: “Yes” | *Not collected in BIS* |
| Method to clean in the house | Mop/vacuum | Q: What is your main method for cleaning dust/ dirt from your floors?<br>R: “Vacuum”; “Other (“Mop”)” | Q: How do you usually clean floor in your main living area (e.g. living room)?<br>R: “Vacuuming”; “Mopping” |
| | Sweep | Q: What is your main method for cleaning dust/ dirt from your floors?<br>R: “Sweep” | Q: How do you usually clean floor in your main living area (e.g. living room)?<br>R: “Sweeping”; “Dusting” |
| Method to clean surfaces | Wet cloth | Q: What is your main method for removing dust from surfaces?<br>R “Wet cloth” | *Not collected in BIS* |
| | Dry cloth/dusting | Q: What is your main method for removing dust from surfaces?<br>R “Dry cloth”; “Dry microfibre cloth”; “Dusting wand” | *Not collected in BIS* |
| Frequency of cleaning floors | Continuous | Q: How many times per week do you clean the dust/dirt from floors? | *Not collected in BIS* |
| Frequency of cleaning surfaces | Continuous | Q: How many times per week do you clean dust/dirt from surfaces? | *Not collected in BIS* |
| Fabric flooring in restroom | No | *Not collected in ELLF* | Q: What kind of floor coverings do you have indoors in your restroom?<br>R: “Tile”; “Wood”; “Linoleum”; “Concrete”; “Slate” |
| | Yes | *Not collected in ELLF* | Q: What kind of floor coverings do you have indoors in your restroom?<br>R1: “Carpet”; “Rug”<br>R2: Have both fabric and non-fabric flooring responses |
| Fabric flooring in kitchen | No | *Not collected in ELLF* | Q: What kind of floor coverings do you have indoors in your kitchen?<br>R: “Tile”; “Wood”; “Linoleum”; “Concrete”; “Slate” |
| | Yes | *Not collected in ELLF* | Q: What kind of floor coverings do you have indoors in your kitchen?<br>R1: “Carpet”; “Rug”<br>R2: Have both fabric and non-fabric flooring responses |
| Fabric flooring in parent’s room | No | *Not collected in ELLF* | Q: What kind of floor coverings do you have indoors in your room?<br>R: “Tile”; “Wood”; “Linoleum”; “Concrete”; “Slate” |
| | Yes | *Not collected in ELLF* | Q: What kind of floor coverings do you have indoors in your room?<br>R1: “Carpet”; “Rug”<br>R2: Have both fabric and non-fabric flooring responses |
| Fabric flooring in baby’s room | No | *Not collected in ELLF* | Q: What kind of floor coverings do you have indoors in your baby’s room?<br>R: “Tile”; “Wood”; “Linoleum”; “Concrete”; “Slate” |

| | | | |
|---|---|---|---|
| | Yes | *Not collected in ELLF* | Q: What kind of floor coverings do you have indoors in your baby's room?<br>R1: "Carpet"; "Rug"<br>R2: Have both fabric and non-fabric flooring responses |
| Total cigarette per day | Continuous | *Not collected in ELLF* | How many cigarettes and/or any tobacco products do you and your partner smoke on average per day? |
| Frequency of cleaning floor in living area | Continuous | *Not collected in ELLF* | Q: How do you usually clean the floor in your main living area?<br>R: "Less than once a month = 1"; "1-3 times a month = 2"; "Once a week = 3"; "A few times a week = 4"; "Every day = 5" |
| Frequency of cleaning floor in child's room | Continuous | *Not collected in ELLF* | Q: How often do you clean the floor in your baby's bedroom?<br>R: "Less than once a month = 1"; "1-3 times a month = 2"; "Once a week = 3"; "A few times a week = 4"; "Every day = 5" |

**Results**

**Random forest models**

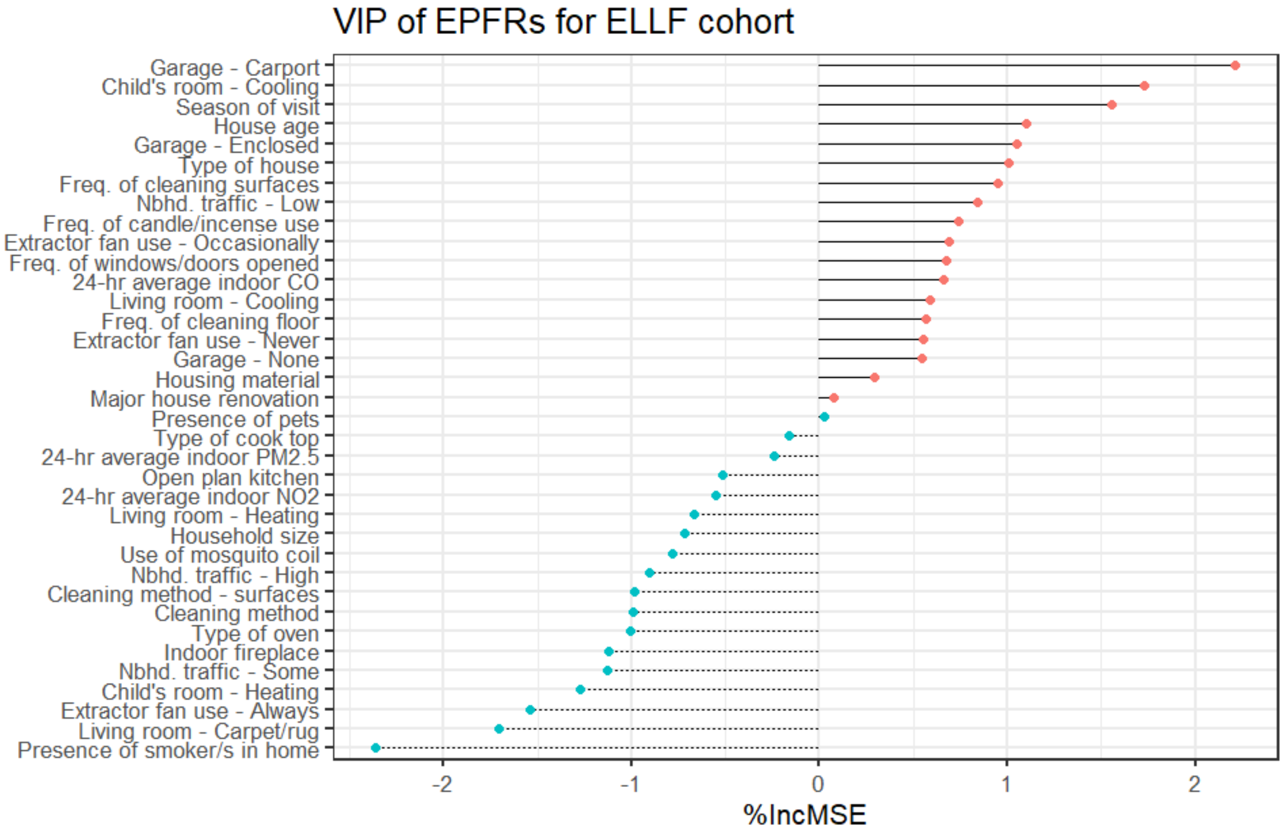


Figure S2 Variable importance plot of most important (red point) to least important household characteristic (blue point) of EPFR concentration in ELLF cohort.

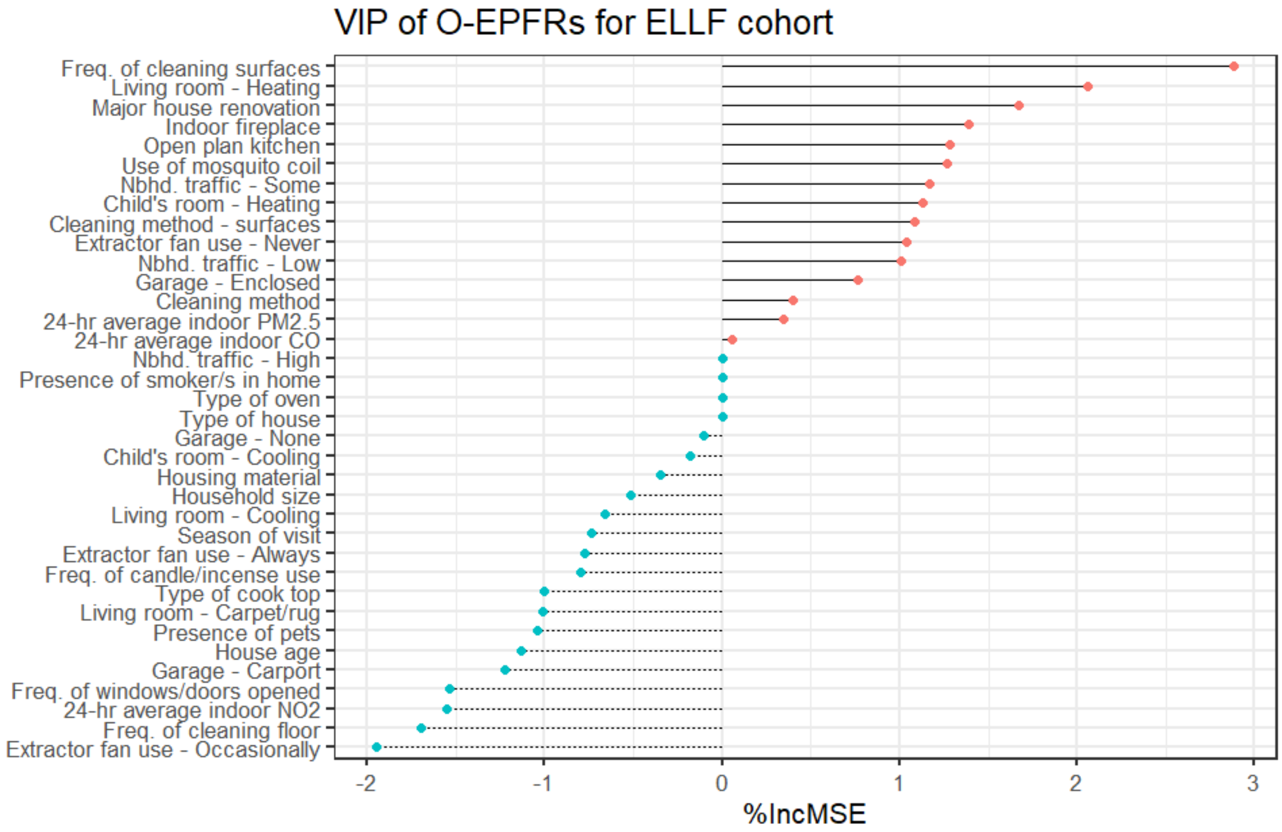


Figure S3 Variable importance plot of most important (red point) to least important (blue point) household characteristic of EPFR oxygen-weighted concentration in ELLF cohort.

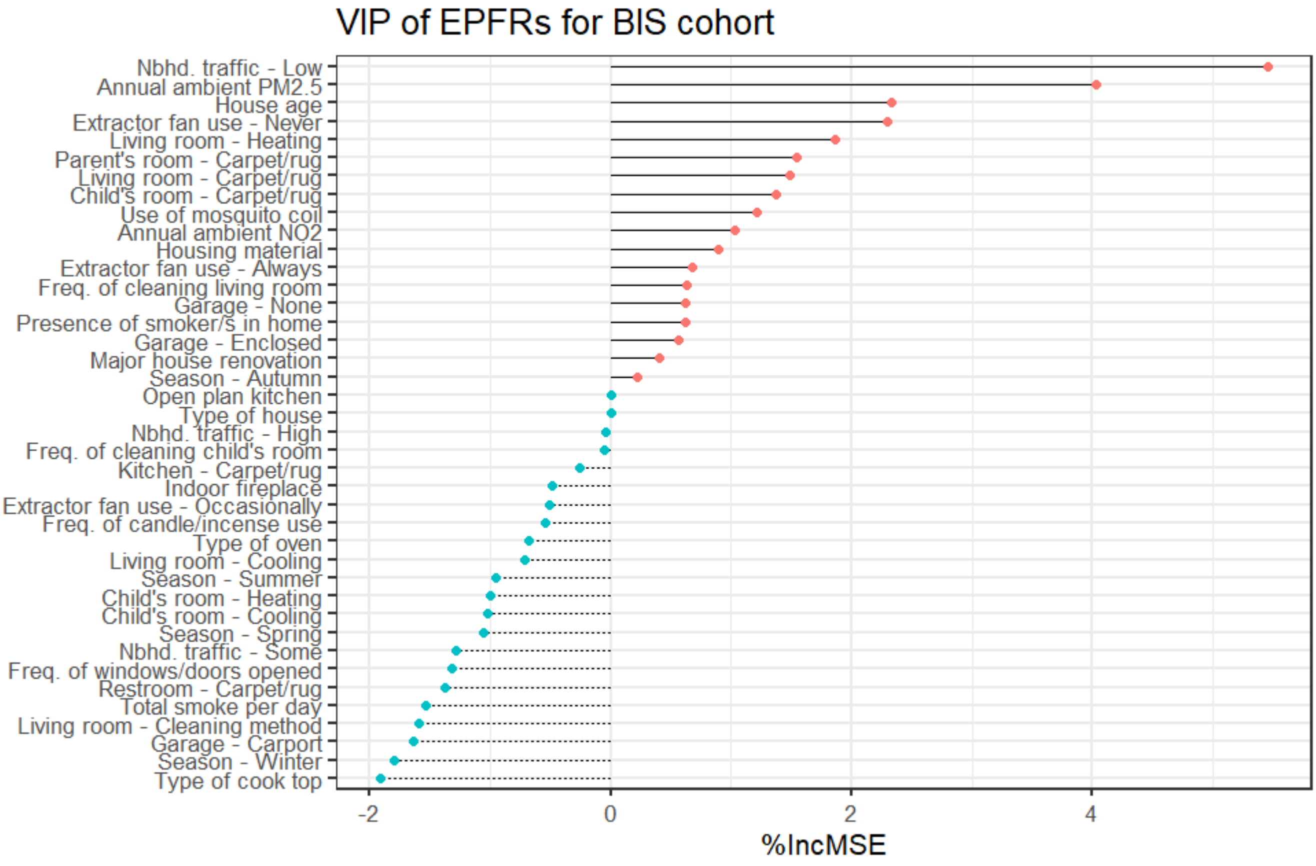


Figure S4 Variable importance plot of most important (red point) to least important (blue point) household characteristic of EPFR concentration in BIS cohort.

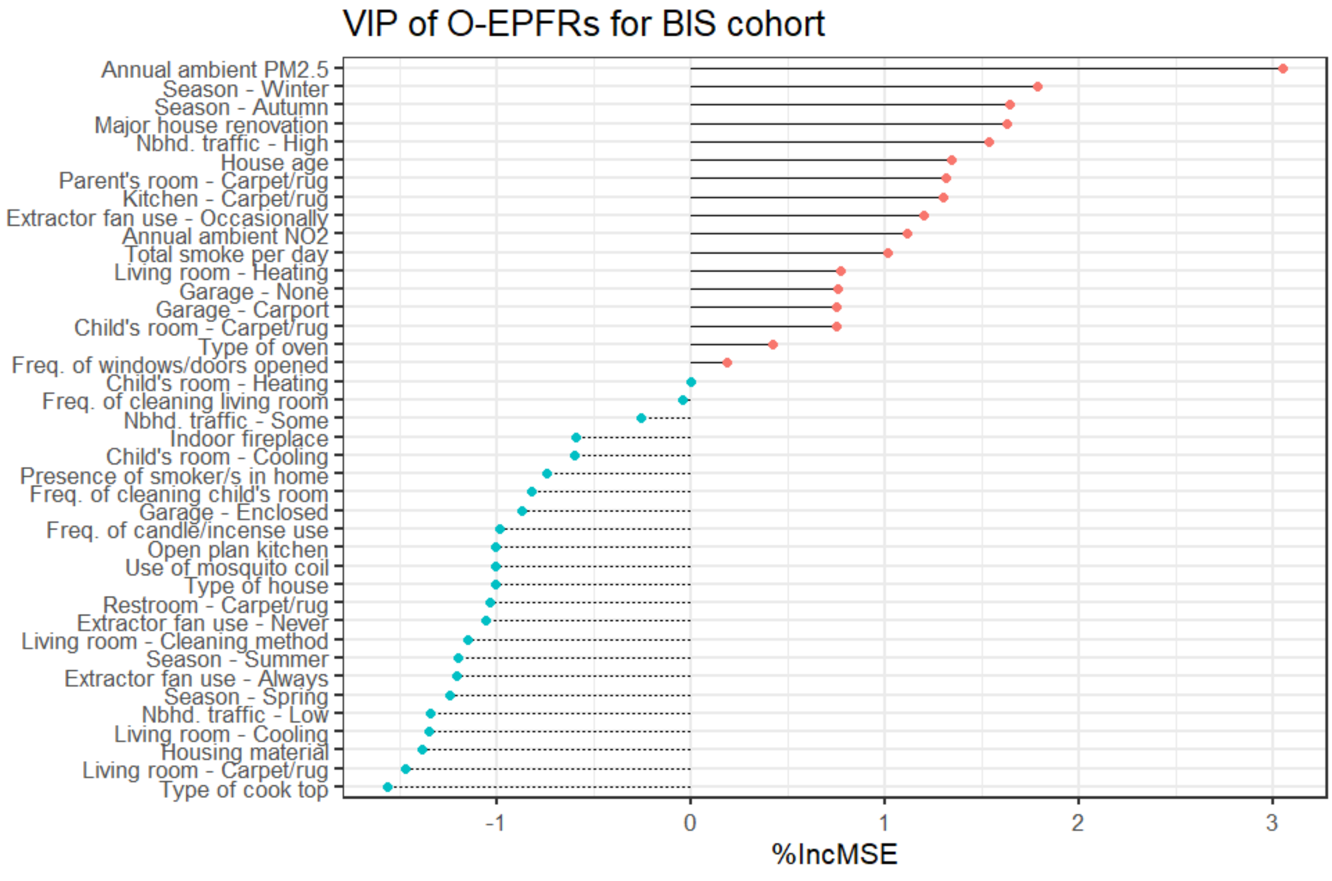


Figure S5 Variable importance plot of most important (red point) to least important (blue point) household characteristic of EPFR oxygen-weighted concentration in BIS cohort.

Table S2 Univariate analysis (Spearman's rho correlation coefficient) of each household variables found important in each random forest model for both ELLF and BIS cohorts.

| Important household variable | Spearman's rho | p-value |
|---|---|---|
| **EPFR concentration in ELLF cohort** | | |
| Carport garage | 0.08 | 0.596 |
| Dirty cooling in child's room | 0.25 | 0.098 |
| Winter season | 0.44 | 0.002 |
| *House age | -0.23 | 0.129 |
| Enclosed garage | 0.16 | 0.301 |
| Flat/unit/apartment | -0.04 | 0.809 |
| *Freq. of cleaning surfaces | -0.02 | 0.871 |
| Low neighbourhood traffic | -0.11 | 0.476 |
| *Freq. of candle/incense use | -0.12 | 0.420 |
| Occ. use of extractor fan | 0.18 | 0.225 |
| *Freq. of windows/doors opened | -0.27 | 0.074 |
| *24-hr average indoor CO | 0.09 | 0.556 |
| Dirty cooling in living room | 0.20 | 0.192 |
| *Freq. of cleaning floors | -0.04 | 0.781 |
| Never use extractor fan | 0.21 | 0.170 |
| No garage | -0.26 | 0.081 |
| W.board housing material | 0.07 | 0.623 |
| Major house renovation | 0.20 | 0.194 |
| **EPFR oxygen-weighted concentration in ELLF cohort** | | |
| *Freq. of cleaning surfaces | -0.34 | 0.048 |
| Dirty heating in living room | 0.35 | 0.037 |
| Major house renovation | 0.18 | 0.300 |
| Indoor fireplace | 0.24 | 0.165 |
| Open plan kitchen | -0.17 | 0.321 |
| Use of mosquito coil | 0.37 | 0.027 |
| Some neighbourhood traffic | 0.18 | 0.293 |
| Clean heating in child's room | -0.10 | 0.581 |
| Clean surfaces by dry cloth | 0.20 | 0.254 |
| Never use extractor fan | 0.15 | 0.374 |
| Low neighbourhood traffic | -0.14 | 0.431 |
| Enclosed garage | 0.20 | 0.243 |
| Clean floors by sweeping | 0.01 | 0.960 |
| *24-hr average indoor PM2.5 | 0.24 | 0.169 |
| *24-hr average indoor CO | 0.01 | 0.970 |
| **EPFR concentration in BIS cohort** | | |
| Low neighbourhood traffic | -0.21 | 0.016 |
| *Annual ambient PM2.5 | 0.18 | 0.041 |
| *House age | 0.26 | 0.002 |
| Never use extractor fan | 0.19 | 0.027 |
| Dirty heating in living area | 0.09 | 0.322 |

| | | |
|---|---|---|
| Carpet/rug in parent's room | 0.13 | 0.132 |
| Carpet/rug in living area | 0.19 | 0.031 |
| Carpet/rug in child's room | 0 | 0.974 |
| Use of mosquito coil | -0.13 | 0.141 |
| *Annual ambient NO2 | 0.11 | 0.192 |
| W.board housing material | 0.24 | 0.005 |
| Always use extractor fan | -0.01 | 0.916 |
| *Freq. of cleaning in living area | 0.04 | 0.681 |
| No garage | 0.03 | 0.690 |
| Presence of smoker/s | 0.12 | 0.166 |
| Enclosed garage | -0.1 | 0.226 |
| Major house renovation | -0.07 | 0.411 |
| Autumn season | -0.04 | 0.653 |
| **EPFR oxygen-weighted concentration in BIS cohort** | | |
| *Annual ambient PM2.5 | 0.24 | 0.012 |
| Winter season | -0.03 | 0.771 |
| Autumn season | 0.01 | 0.942 |
| Major house renovation | -0.14 | 0.164 |
| High neighbourhood traffic | 0.19 | 0.049 |
| *House age | 0.22 | 0.021 |
| Carpet/rug in parent's room | 0.03 | 0.757 |
| Carpet/rug in kitchen | 0.17 | 0.078 |
| Occ use extractor fan | -0.1 | 0.325 |
| *Annual ambient NO2 | 0.14 | 0.166 |
| Total cigarette smoke per day | 0.13 | 0.202 |
| Dirty heating in living area | 0.05 | 0.629 |
| No garage | -0.01 | 0.913 |
| Carport | 0.02 | 0.860 |
| Carpet/rug in child's room | -0.09 | 0.369 |
| Gas oven | 0.10 | 0.305 |
| Freq. of windows/doors opened | 0.00 | 0.998 |

**Sensitivity analysis**

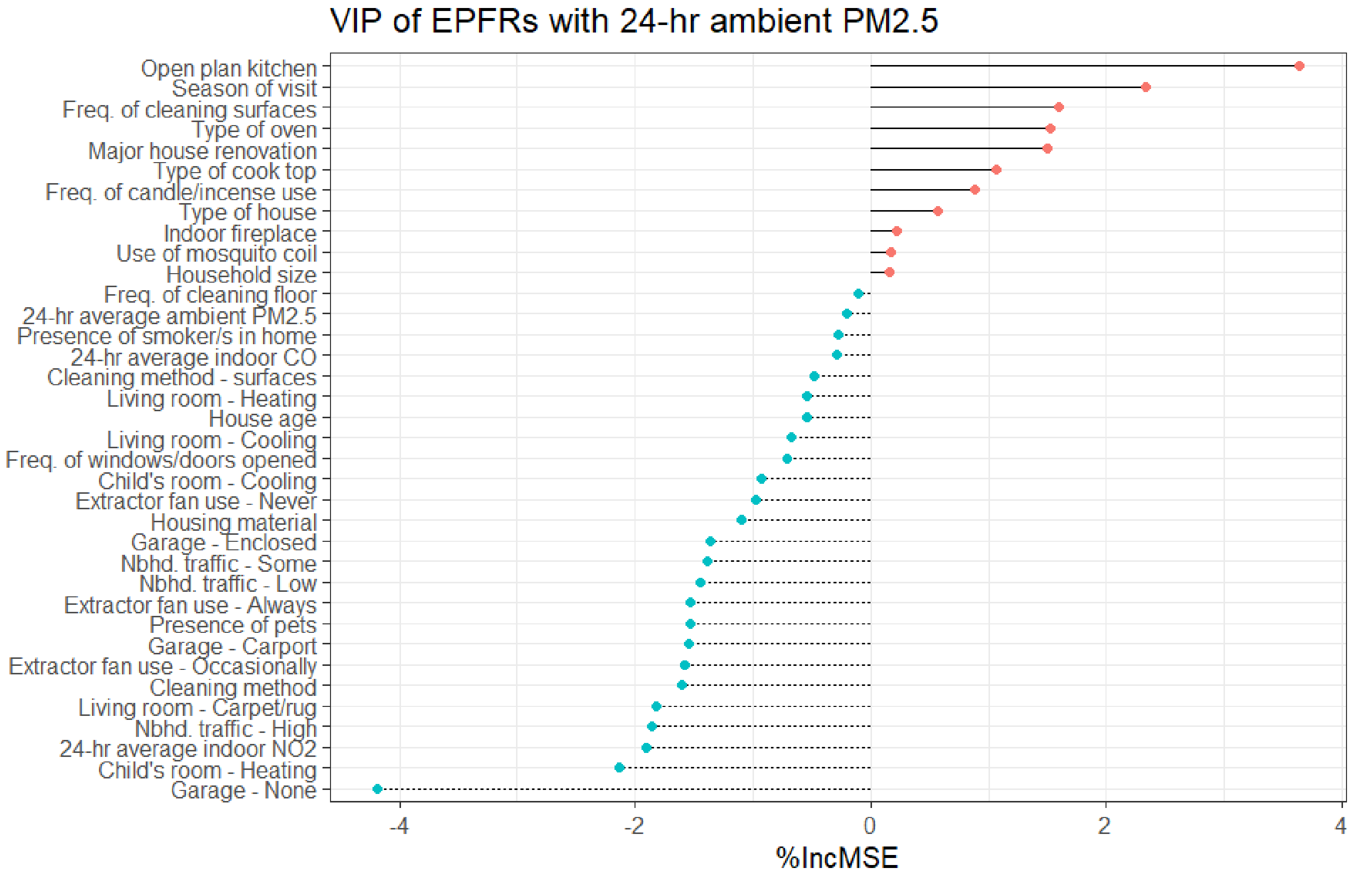


Figure S6 Variable importance plot of ambient $PM_{2.5}$ from most important (red point) to least important (blue point) household characteristic of EPFR concentration in ELLF cohort.

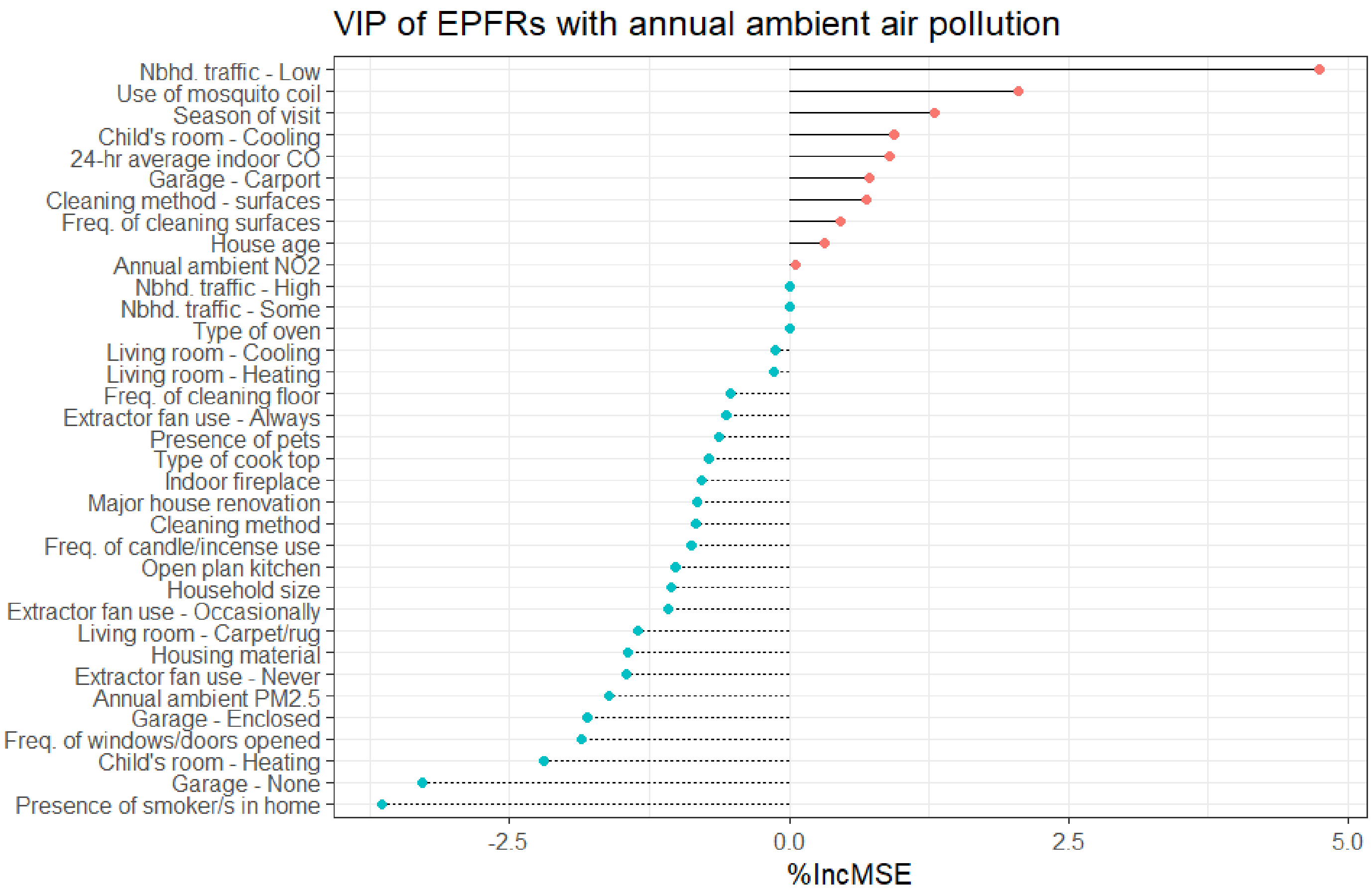


Figure S7 Variable importance plot of annual ambient air pollution from most important (red point) to least important (blue point) household characteristic of EPFR concentration in ELLF cohort.

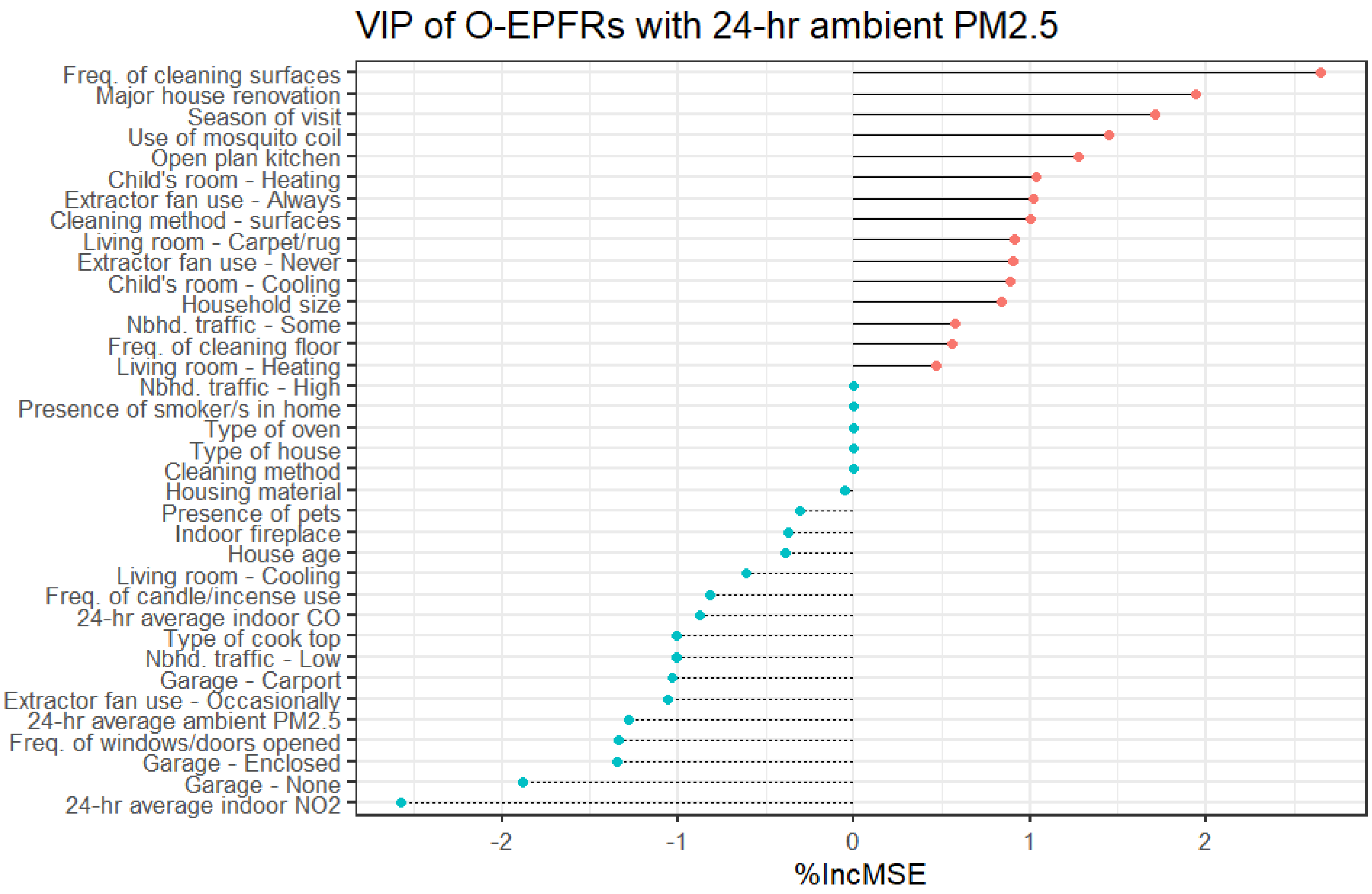


Figure S8 Variable importance plot of ambient $PM_{2.5}$ from most important (red point) to least important (blue point) household characteristic of EPFR oxygen-weighted concentration in ELLF cohort.

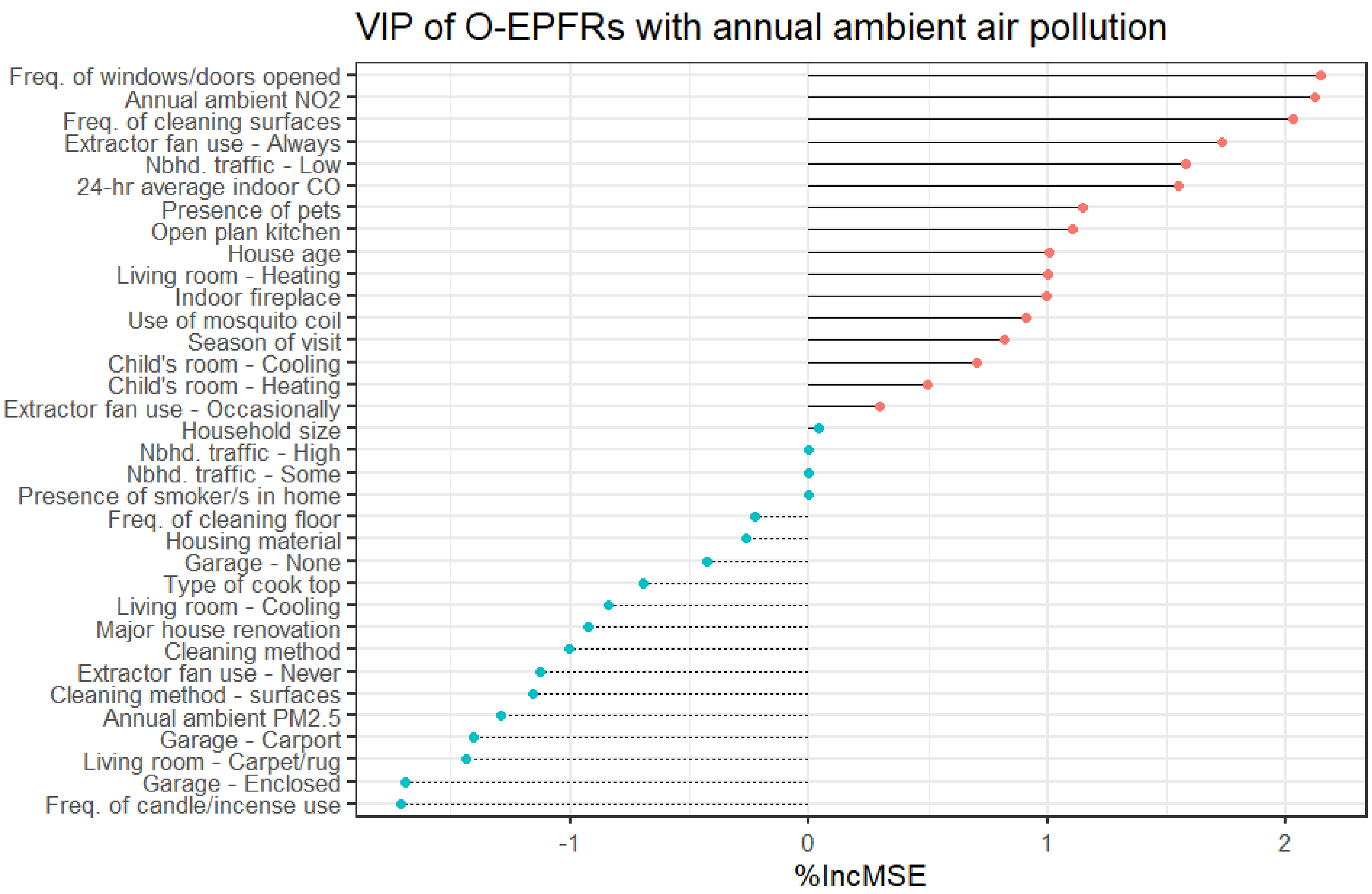


Figure S9 Variable importance plot of annual ambient air pollution from most important (red point) to least important (blue point) household characteristic of EPFR oxygen-weighted concentration in ELLF cohort.
